# An epidemiological and intervention assessment of the malaria epidemic in Bolívar, Venezuela: a modelling study

**DOI:** 10.1101/2022.04.19.22274042

**Authors:** John H. Huber, Luis F. Chaves, Amir S. Siraj, Jorge E. Moreno, Maria Eugenia Guevara, Maria Villegas, Leonor Pocaterra, Leopoldo Villegas, T. Alex Perkins

## Abstract

**Background:** Venezuela has experienced an explosive resurgence in *Plasmodium falciparum* and *Plasmodium vivax* malaria incidence over the last decade due to various social, political, and economic factors. To ensure national and regional progress towards malaria elimination, there is an urgent need to better understand the epidemiological dynamics of this malaria outbreak at its epicenter in the southern state of Bolívar and to identify the sets of interventions that may be necessary to reduce transmission and incidence.

**Methods:** We fitted transmission models of *P. falciparum* and *P. vivax* to weekly incidence data in Bolívar, Venezuela during 2000-2018. We estimated the magnitude of local transmission for both *Plasmodium* spp. and inferred the contribution of relapses and reinfections to *P. vivax* incidence in the region. Compared to a business-as-usual scenario, we projected the impact of different interventions on *Plasmodium* spp. incidence during the period 2021-2023.

**Findings:** We estimated that 63·7 – 73·3% of all *P. vivax* infections in Bolívar are relapses, leading to as many as 51,800 observed relapses misclassified per year as reinfections in the routine surveillance data. Our estimates suggest that the reproduction number remains close to one for both *Plasmodium* spp., pointing towards the feasibility of control. Long-lasting insecticidal nets (LLINs) were projected to cause greater proportional reductions in *P. falciparum* incidence than *P. vivax* incidence, and mass drug administration (MDA) with an 8-aminoquinoline and a blood-stage partner drug was projected to cause the greatest reduction in *P. vivax* incidence, provided that adherence rates were high.

**Interpretation:** Control of the malaria outbreak in Southeastern Venezuela is feasible, should appropriate resources to support surveillance and control be brought to bear. Coupling the distribution of LLINs and a focal MDA with an 8-aminoquinoline and a blood-stage partner drug may lead to the greatest reduction in malaria incidence.

**Funding:** National Science Foundation; University of Notre Dame; National Institute of General Medical Sciences (grant number 1R35GM143029-01 to TAP);

**RESEARCH IN CONTEXT:** *Evidence before the study:* We searched PubMed, bioRxiv, and medRxiv for articles in English published on or before May 25^th^, 2021 using the following keywords: “Venezuela”, “malaria”, AND “model*”. Previous studies have applied statistical models to characterize the relationship between malaria incidence and climate in Venezuela, concluding that the reproduction number is low and suggesting the feasibility of control. A study fitting a mechanistic transmission model to epidemiological data to allow for projecting the impact of alternative approaches to control has not been performed.

*Added value of the study:* We fitted *Plasmodium falciparum* and *Plasmodium vivax* transmission models to 20 years of weekly incidence data to estimate the transmission of both *Plasmodium* spp. and characterize the contribution of relapses and reinfections to *P. vivax* incidence in Bolívar, Venezuela. We also projected the likely impact of interventions in the region under alternative scenarios about control.

*Implications of the available evidence:* The burden of *Plasmodium vivax* relapses in Bolívar is underestimated from routine surveillance data, so control interventions must target the hypnozoite reservoir in the region. Mass drug administration (MDA) is projected to be impactful for both *Plasmodium* spp., though tradeoffs between coverage and adherence suggest that a focal MDA with an 8-aminoquinoline and a blood-stage partner drug may yield the greatest impact.

## INTRODUCTION

Venezuela has witnessed a massive resurgence in *Plasmodium falciparum* and *Plasmodium vivax* malaria incidence over the last two decades and now accounts for more than 70% of all malaria-related deaths in Latin America.^1^ This public health crisis is centered in the southern state of Bolívar and is driven by multiple factors, including a fragile economy,^2^ increased migration,^3–5^ decreased malaria expenditure,^1^ and an expansion of mining-related activities in malaria-receptive areas.^6–8^

Efforts to address the malaria resurgence in Venezuela are urgently needed to ensure that the WHO Region of the Americas meets the targets established by the Global Technical Strategy for Malaria, 2016-2030.^1^ Recently, the Global Fund to Fight AIDS, Tuberculosis, and Malaria approved Venezuela to receive additional funding to expand malaria control efforts.^9^ To achieve the greatest impact, the selection of interventions as part of this expansion should be guided by the local epidemiological dynamics in Bolívar, the epicenter of the malaria resurgence. In an area with co-circulation of *P. vivax* and *P. falciparum*, the immediate impacts of interventions, such as long-lasting insecticidal nets (LLINs) and mass drug administration (MDA), on malaria incidence depend upon the extent to which *P. vivax* infections are reinfections caused by mosquito biting or relapses caused by hypnozoite activation. If most *P. vivax* infections are reinfections, then interventions that can achieve reductions in *P. falciparum* burden, such as LLINs, will likely be effective in reducing *P. vivax* burden, too. By contrast, if most *P. vivax* infections are relapses, then interventions that target the hypnozoite reservoir, such as MDA with an 8-aminoquinoline, may be needed.

Disentangling the contributions of reinfections and relapses to *P. vivax* burden in Bolívar using epidemiological data alone is challenging, because reinfections and relapses cannot be easily distinguished using standard diagnostic criteria. Fitting mathematical models—that explicitly account for both transmission dynamics and the possibility of diagnostic errors—to epidemiological data is an advantageous approach to both characterize the nature of the local malaria burden and inform the best approaches to control.^10^

In this modelling study, we fitted mechanistic models of *P. falciparum* and *P. vivax* transmission to epidemiological data collected in Bolívar, Venezuela during 2000-2018. We first analyzed changes in the epidemiological dynamics of *P. falciparum* and *P. vivax* over the same time period using the fitted models and then projected the likely impact of interventions in the region under alternative scenarios about control.

## METHODS

### Malaria Transmission Models

We developed deterministic compartment models to characterize the transmission dynamics of *P. falciparum* and *P. vivax* malaria in Bolívar, Venezuela. To account for changes in transmission over time, the models used a time-varying form of the reproduction number derived from first principles of transmission, which calculates the expected number of secondary infections arising from each infected individual.^11^ By treating the reproduction number as time-varying, we accounted for the effects of temporal variation in mosquito and parasite traits on transmission^12–14^ and were able to model the effects of interventions on each component of the transmission cycle.^10,15^

In the *P. falciparum* model, individuals are initially susceptible and are then infected according to the time-varying reproduction number. Following infection, a proportion of individuals seek treatment with artemisinin combination therapy (ACT) in a health clinic, and the remaining proportion of individuals are untreated and clear their infection naturally. In Venezuela, the first-line ACT used is Artemether-Lumefantrine (AL). To model ACT adherence, we assumed that a proportion of treated individuals adhere fully to ACT and clear the *P. falciparum* infection. The remaining proportion of treated individuals were non-adherent to ACT and experienced a recrudescing infection. The duration of a *P. falciparum* infection depended upon whether an individual received ACT and was generated using a within-host model from Johnston *et al*.^13,14^ Following clearance of the infection, individuals enter into a recovered state, where they experience a period of temporary immunity that is exponentially distributed.

The *P. vivax* model is structurally similar to the *P. falciparum* transmission model, though it accounts for the latent, liver-stage *P. vivax* parasites, known as hypnozoites, which can cause relapsing infections. Initially, individuals are susceptible and free from hypnozoites and are infected according to the time-varying reproduction number. We assumed that each *P. vivax* infection resulted in both a primary bloodstream infection and a latent liver infection with hypnozoites. As was modeled in the *P. falciparum* transmission model, a proportion of individuals seek treatment with chloroquine (CQ) in a health clinic, and a proportion of treated individuals also receive radical cure with primaquine (PQ). Depending upon the treatment regimen, treatment of a *P. vivax* infection could result in one of two outcomes: (1) complete clearance of the primary bloodstream and latent liver infections or (2) clearance of the primary bloodstream infection only. Untreated individuals were assumed to clear only the primary bloodstream infection. Following clearance of the bloodstream infection, individuals with hypnozoites could relapse or become re-infected. Upon clearance of both primary bloodstream and latent liver infections, all individuals enter into a recovered state, where they experience a period of temporary immunity that is exponentially distributed. Following loss of immunity, individuals are once again susceptible and can be re-infected.

In both transmission models, we accounted for changes in the population through births and deaths.^16^ We did not model movement within Bolívar, nor did we address heterogeneity in transmission risk. Further details of the *P. falciparum* and *P. vivax* transmission models are provided in the appendix.

### Intervention Models

We modeled the impact of long-lasting insecticidal nets (LLIN) and mass drug administration (MDA) compared to the existing standard of care for clinical episodes of *P. falciparum* and *P. vivax*.

An increase in LLIN coverage was assumed to impact *P. falciparum* and *P. vivax* transmission through a reduction in mosquito biting, an increase in mosquito mortality, and a corresponding reduction in the mosquito population.^15^ We modeled reductions in LLIN usage and insecticidal activity over time and assumed that the mean durations of LLIN usage and insecticidal activity were 84·2 and 130 weeks, respectively.^10^ A supplementary analysis was performed to evaluate the sensitivity of the projected impact to the assumed parameter values.

The effect of MDA on *P. falciparum* and *P. vivax* transmission depended upon the antimalarial drugs included. Treatment with ACT was assumed to clear both *P. falciparum* and *P. vivax* bloodstream infections and, by default, provided three weeks of prophylaxis. We assumed that treatment with CQ cleared *P. vivax* bloodstream infections and, by default, provided two weeks of prophylaxis. Only radical cure with PQ could clear the latent liver-stage *P. vivax* infection, and adherence to PQ was assumed to be 100% at baseline. For all MDA campaigns, we calculated the effective coverage from a target coverage by accounting for contraindications due to pregnancy and G6PD status. A complete description of all interventions can be found in the appendix.

### Model Fitting

We used weekly incidence data during 2000-2018 and cumulative incidence data during 2018-2019 reported by the Venezuelan Ministry of Health as well as annual prevalence estimates generated by the Malaria Atlas Project during 2000-2017.^17^ To obtain a counterfactual against which our impact projections were evaluated, we produced forward simulations of the transmission models during 2021-2023.

We fitted our *P*. falciparum and *P. vivax* transmission models to the epidemiological data using a Bayesian framework. To account for spatial heterogeneity within Bolívar that could affect impact projections, we similarly re-fitted our transmission models to data from the Sifontes municipality, the epicenter of the malaria epidemic within Bolívar. A full description of the model-fitting procedure, as well as the parameters estimated during model fitting, are provided in the appendix.

### Scenario Analyses

We projected the impact of alternative intervention packages on *P. falciparum* and *P. vivax* transmission in Bolívar. The intervention packages considered beyond the existing standard of care were distribution of LLINs and the implementation of MDA.

All intervention packages were assumed to be implemented at the start of 2021. For LLIN distribution, we varied the coverage from 10-90% in increments of 10%. For MDA, we varied the coverage from 50-80% in increments of 5%. We also varied the choice of antimalarial drugs and the assumed level of adherence. These settings were assumed to apply consistently throughout the intervention period.

We calculated the absolute reduction and the percent reduction in *P. falciparum* and *P. vivax* incidence during 2021-2023 for each intervention package relative to a business-as-usual scenario. Full details of the implementation of the intervention packages are provided in the appendix.

## RESULTS

The fitted transmission models captured the long-term changes and seasonal fluctuations therein in *P. falciparum* and *P. vivax* incidence in Bolívar (Fig. 1). We estimated that *P. falciparum* incidence increased over the period 2000-2018 and then decreased during 2019-2020 and that *P. vivax* incidence increased over the period 2000-2020. The estimated divergent trajectories in *P. falciparum* and *P. vivax* incidence matched the stable cumulative *Plasmodium* spp. incidence from 2018 to 2019, the time period for which complete weekly incidence data were not available (Fig. S10A). Furthermore, our estimates of annual *P. vivax* prevalence during 2000-2017 were consistent with the increase in predicted geospatial prevalence estimates to which the transmission model was fitted (Fig. S10B).

**Figure 1.**
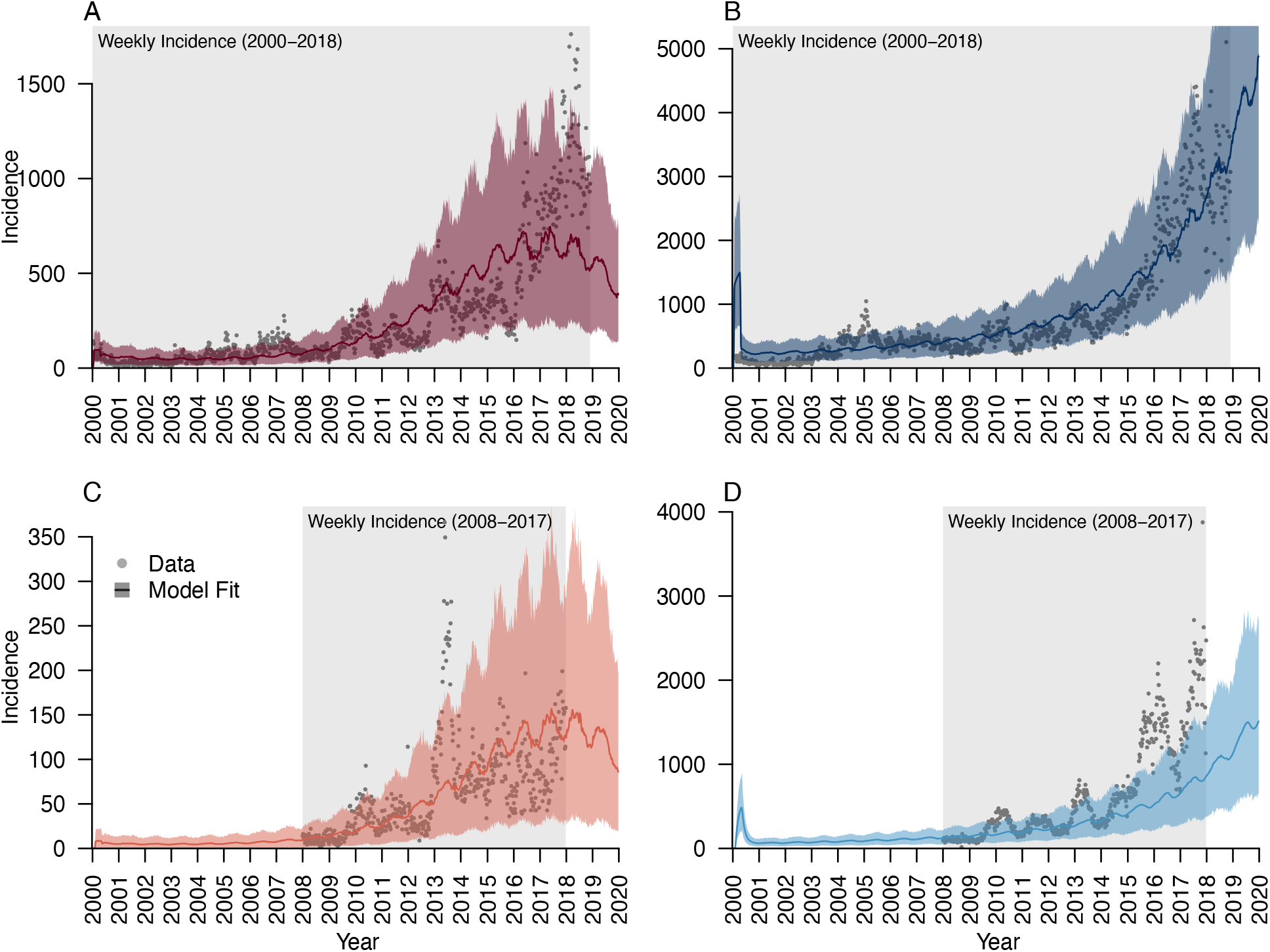
Comparison of fitted transmission model to weekly epidemiological data. The fitted P. falciparum and P. vivax transmission models are compared to the weekly incidence data for (A) reported P. falciparum new infections, (B) reported P. vivax new infections, (C) reported P. falciparum recrudescing infections, and (D) reported P. vivax relapses. The shaded gray area represents the period for which weekly incidence data was available, and each gray point is the reported incidence in a given week. The colored line is the median posterior estimate from the fitted transmission model, and the colored shaded region is the 95% posterior prediction interval.

The epidemiological dynamics of the fitted *P. vivax* transmission model indicated that the epidemiological data may have underestimated the extent of relapsing infections in Bolívar. On the basis of the epidemiological data alone, the annual proportion of *P. vivax* cases that were classified as relapses ranged from 24·3% in 2008 to 46·6% in 2015, suggesting that most *P. vivax* infections were caused by local transmission, not relapses. However, the fitted transmission model estimated that the annual proportion of all *P. vivax* infections that were relapses ranged from 67·6% (67·3 – 67·8%) to 71·4% (71·2 – 71·6%) over the same time period (Fig. 2C). This difference between the epidemiological data and the estimated transmission dynamics was explained by the large number of relapses that we inferred to have been misclassified as new infections. We estimated that in 2018 the annual number of relapses that were misclassified as new infections was 32,500 (95% PPI: 14,500 – 61,400), while the annual number of new infections that were misclassified as relapses was only 52 (0 – 189) (Fig. 2B).

**Figure 2.**
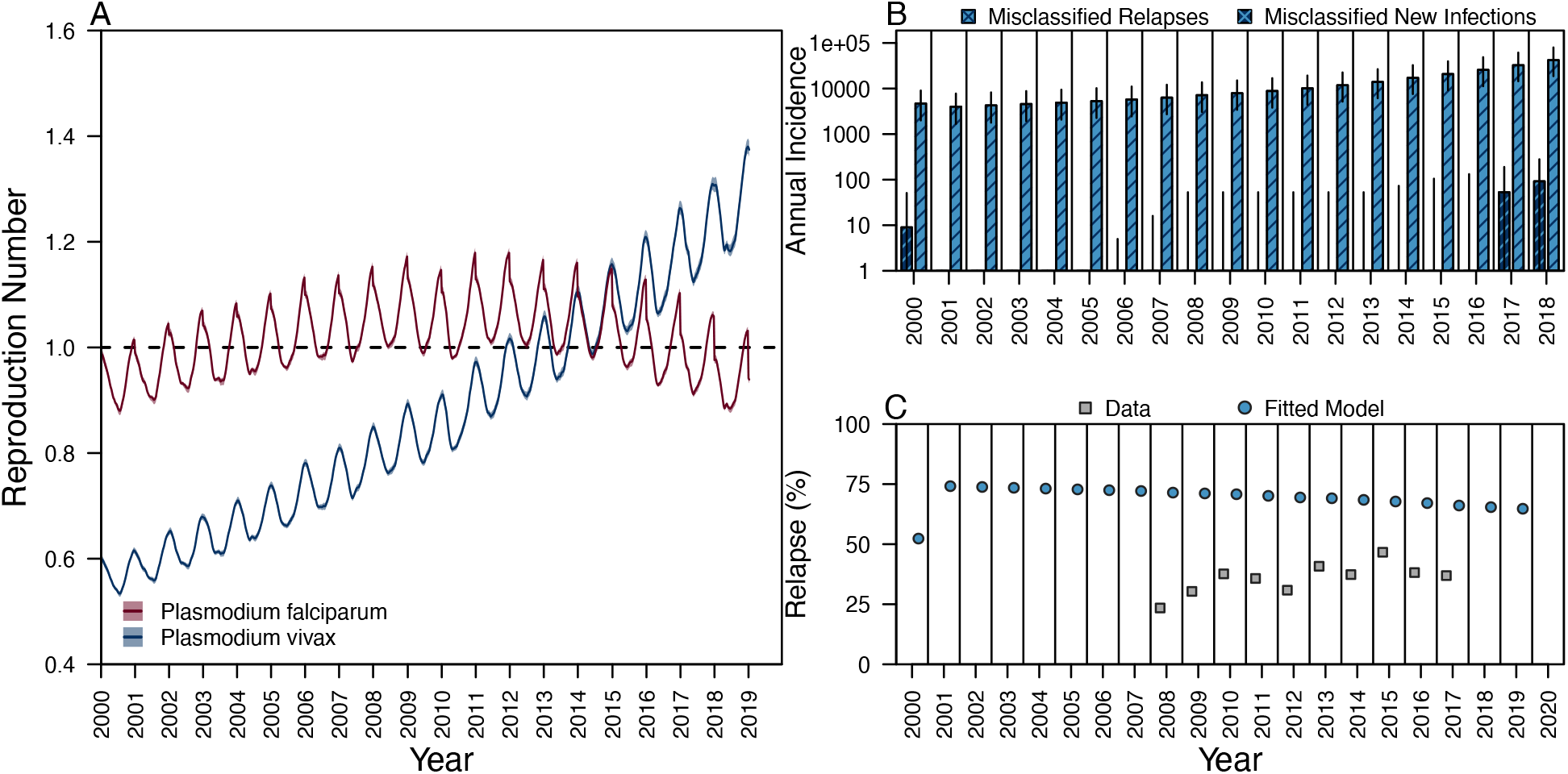
Transmission and epidemiological dynamics in Bolívar, Venezuela. (A) The posterior median (line) and 95% credible interval (shaded region) of the time-varying reproduction number are shown for P. falciparum (red) and P. vivax (blue). (B) The annual incidence of P. vivax relapses that are misclassified as new infections (light blue) and P. vivax new infections that are misclassified as relapses (dark blue) are shown during 2000-2018. Segments denote the 95% posterior prediction intervals. (C) The annual percentage of all P. vivax infections during 2000-2020. Each gray point is the percentage obtained from the epidemiological data (available during 2008-2017), and each blue point is the percentage of all P. vivax infections, both reported and unreported, from the fitted model.

We also examined changes in the transmission dynamics during 2000-2020. We estimated that the reproduction numbers for both *Plasmodium* spp. have remained close to one over the time period considered (Fig. 2A), though seasonal fluctuations were estimated to cause transmission to peak around the second-to-last week of April in each year. That the reproduction numbers have remained close to one indicated the feasibility of control should interventions be implemented. Nevertheless, estimating transmission at the state level may obscure important heterogeneity in transmission risk, and transmission models fitted to the Sifontes municipality, the epicenter of the outbreak in Bolívar, revealed a potentially higher level of transmission of *P. vivax* (Fig. S13).

The data on historical interventions in Bolívar indicated that the coverage of LLINs was relatively low over the period 2000-2018 and likely did not have an effect on transmission over this time period. We projected that the distribution of LLINs occurring at the start of 2021 with a target coverage of 10% would avert 20,200 (18,700 – 21,200) *P. falciparum* infections and 168,000 (167,000 – 170,000) *P. vivax* infections over the time period 2021-2023 (Fig. 3A).

**Figure 3.**
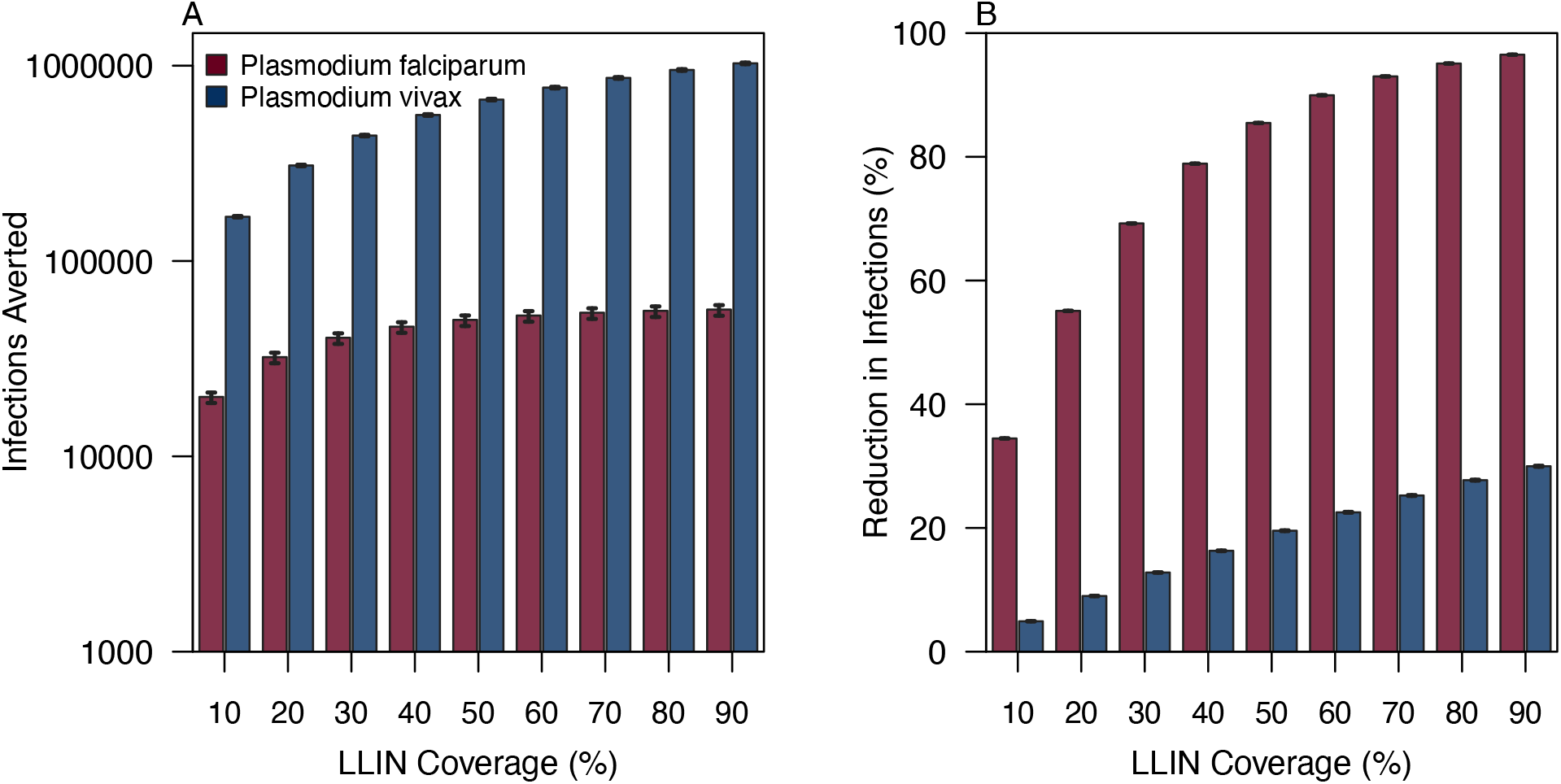
Projected impact of LLIN distribution during 2021-2023. (A) The expected number of P. falciparum (red) and P. vivax (blue) infections averted during 2021-2023 is projected as a function of LLIN coverage. (B) The effect size of LLIN distribution is projected as a function of LLIN coverage. In both plots, the height of the bar denotes the median of the posterior prediction distribution, and the segments are the 95% posterior prediction interval.

Under an alternative scenario with a target coverage of 90%, we projected that the distribution of LLINs would avert 56,400 (52,500 – 59,400) *P. falciparum* infections and 1.03 million (1.02 – 1.04 million) *P. vivax* infections over the time period 2021-2023. When measured as proportional reduction in incidence relative to a business-as-usual scenario, the distribution of LLINs is expected to be more impactful for *P. falciparum* than *P. vivax* at all target coverages considered (Fig. 3B). The diminished projected impact of LLINs for *P. vivax* relative to *P. falciparum* was attributed to the high proportion of relapsing infections (Fig. 2C), which are not prevented by the use of LLINs.

The overall impact of mass drug administration of antimalarial drugs that target blood-stage *Plasmodium* spp. parasites was projected to be more similar across *Plasmodium* spp. than for the distribution of LLINs (Figs. 3B, 4B). At a target coverage of 50%, we projected that MDA of an antimalarial drug providing 14 days of prophylaxis would result in 30·1% (29·9 – 30·4%) and 49·0% (48·8 – 49·3%) reductions in *P. falciparum* and *P. vivax* incidence over 2021-2023, corresponding to respective totals of 17,600 (16,500 – 18,600) and 1,680,000 (1,660,000 – 1,700,000) infections averted. Under an alternative scenario with a target coverage of 80%, we projected that an MDA of an antimalarial drug providing 14 days of prophylaxis would cause 34·7% (34·4 – 34·9%) and 63·7% (63·4 – 63·9%) reductions in *P. falciparum* and *P. vivax* incidence over 2021-2023, corresponding to respective totals of 20,300 (18,900 – 21,400) and 2,180,000 (2,150,000 – 2,210,000) infections averted. Using an antimalarial drug with a longer duration of prophylaxis provided a greater projected impact for *P. falciparum* than *P. vivax*, because most *P. vivax* infections were due to relapse, not reinfection (Fig. 2C). At a target coverage of 50%, the proportional reductions in *P. falciparum* and *P. vivax* infections over 2021-2023 were 38·4% (38·2 – 38·6%) and 49·8% (49·5 – 50·0%) respectively when the duration of prophylaxis was 28 days, compared to 30·1% (29·9 – 30·4%) and 49·0% (48·8 – 49·3%) when the duration of prophylaxis was 14 days.

**Figure 4.**
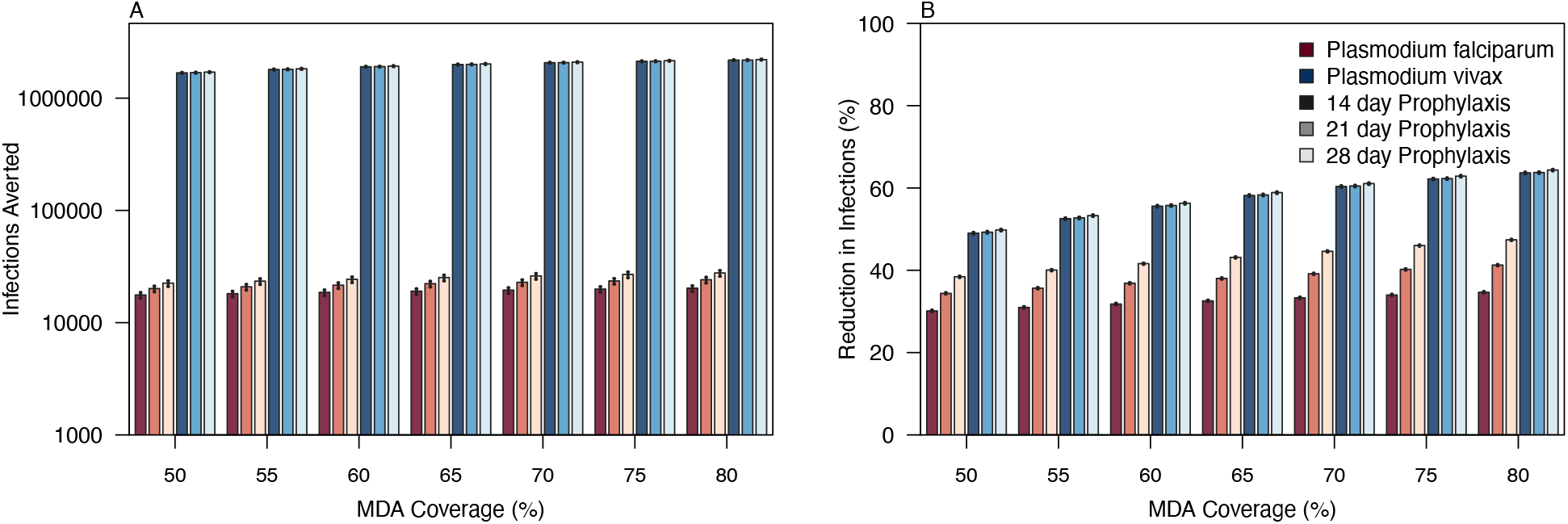
Projected impact of MDA (CQ vs. ACT) during 2021-2023. (A) The expected number of P. falciparum (red) and P. vivax (blue) infections averted during 2021-2023 is projected as a function of MDA coverage for therapeutics with varying durations of prophylaxis. (B) The effect size of each MDA is projected as a function of coverage. Adherence to treatment is assumed to be 100%. In both plots, the height of the bar is the median of the posterior prediction distribution, and the segments are the 95% posterior prediction interval.

Because most *P. vivax* infections were relapses in the fitted model (Fig. 2B & 2C), we next projected the impact of MDA of radical cure, which clears the hypnozoites in the liver that cause relapses. Assuming 100% adherence at a target coverage of 50%, MDA with PQ and a blood-stage, antimalarial drug providing 14 days of prophylaxis was projected to avert 2.13 million (2.10 –2.15 million) *P. vivax* infections during 2021-2023, a reduction of 62·2% (62·0% - 62·4%). By comparison, increasing the target coverage to 80% was projected to avert 2.57 million (2.54 million – 2.60 million) infections, a reduction of 75·0% (74·8% - 75·2%) over the same time period. These impact projections are likely an underestimate, because our model did not account for the gametocidal activity of PQ, which may further reduce *Plasmodium* spp. transmission. Nevertheless, our results underscore the potential impact of radical in MDA campaigns on the hypnozoite reservoir in Bolívar.

Our projected impact of MDA with radical cure assumed, by default, complete adherence to PQ. Primaquine adherence, however, is low given the length of treatment, and measures taken to ensure high rates of adherence may be operationally infeasible at the target coverages considered. We therefore examined how our impact projections varied as a function of MDA coverage and PQ adherence. We found that the expected impact of MDA with CQ + PQ depended upon the assumed PQ adherence. At a target coverage of 50%, the expected reduction in *P. vivax* infections ranged from 49% at 0% PQ adherence to 60% at 100% PQ adherence (Fig. 6B). By contrast, at 50% PQ adherence, increasing the target coverage from 50% to 80% increased the expected reduction in *P. vivax* infections from 55% to 70%. Our results suggest that comparable projected impacts on *P. vivax* can be achieved under both low coverage, high adherence and high coverage, low adherence scenarios. Focal MDA that targets a smaller subset of the population at higher risk of exposure to *P. vivax* and ensures high PQ adherence may be preferred to an intervention strategy that maximizes the target coverage.

**Figure 5.**
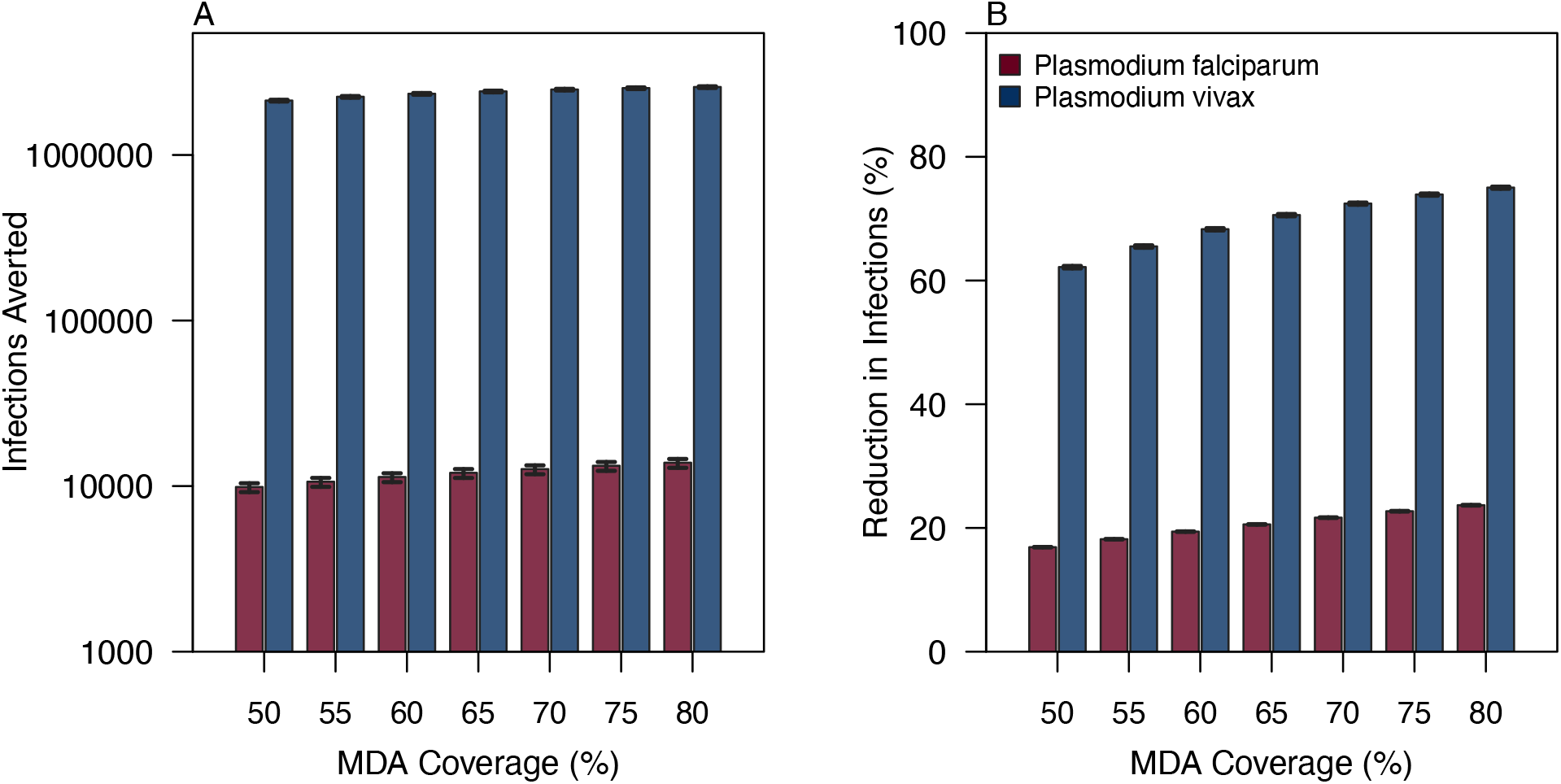
Projected impact of MDA (CQ + PQ) during 2021-2023. (A) The expected number of P. falciparum (red) and P. vivax (blue) infections averted during 2021-2023 is projected as a function of MDA coverage. (B) The effect size of MDA is projected as a function of coverage. Adherence to treatment is assumed to be 100%. In both plots, the height of the bar denotes the median of the posterior prediction distribution, and the segments are the 95% posterior prediction interval.

**Figure 6.**
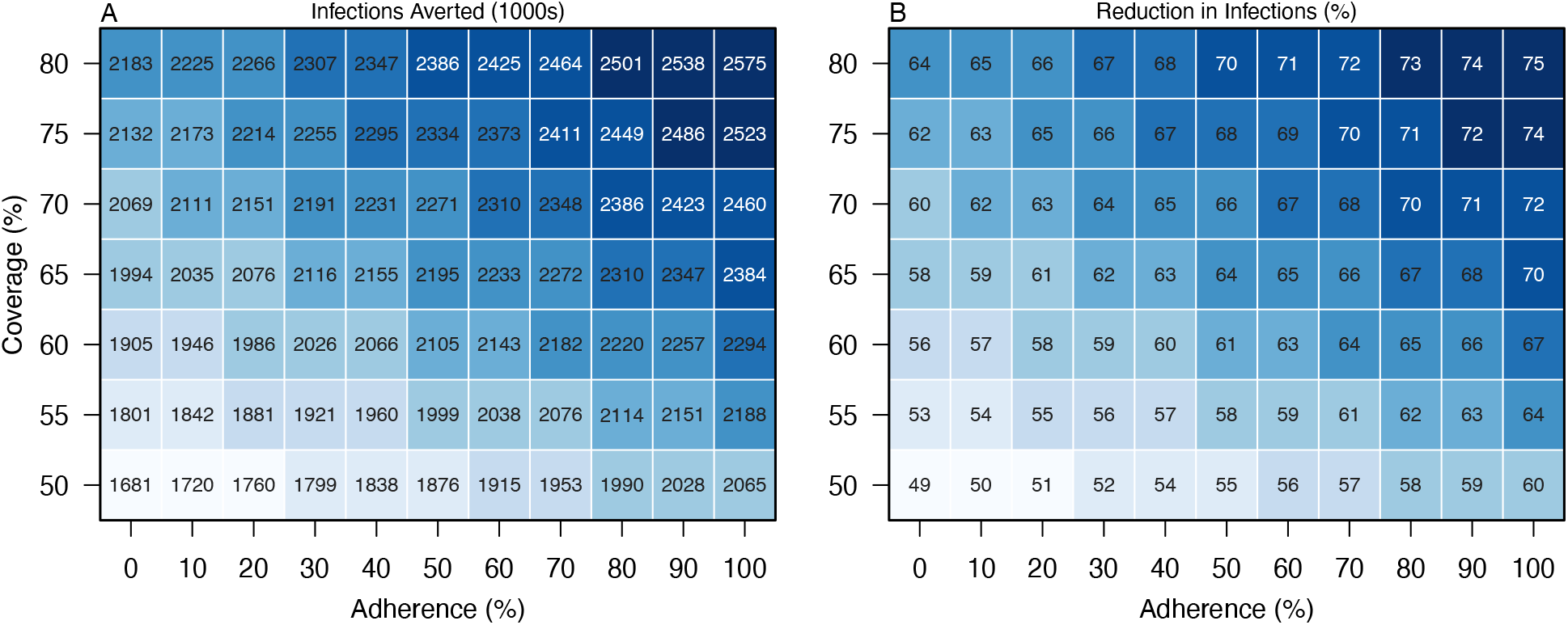
Effect of coverage and adherence on the projected impact of MDA (CQ + PQ) for P. vivax. The expected number of P. vivax infections averted (A) and the effect size (B) across 2021-2023 is presented for each combination of coverage and adherence in the target population. The intensity of the color denotes the median projected impact, where lighter colors denote lower impact and darker colors denote high impact.

## DISCUSSION

Malaria incidence in southeastern Venezuela has increased sharply over the last decade, and control interventions are needed to address this public health crisis.^6,8^ When selecting control interventions to combat malaria resurgence in Bolívar, policy makers must consider the local epidemiological dynamics in addition to logistical and cost constraints. Our model-guided assessment suggests that the use of surveillance data alone provides an incomplete understanding of the epidemiological and transmission dynamics in the region. Our results indicate that surveillance data may have underestimated the relative burden of relapses among *P. vivax* cases as compared to reinfections, due to misclassification arising from narrow case definitions. The case definition of a relapse required clinical presentation within 90 days following a recent clinical episode of *P. vivax*. Thus, *P. vivax* relapses that had an asymptomatic or undetected primary infection or that occurred outside the 90-day window would be misclassified, as would *P. vivax* reinfections that occurred with 90 days of a recent clinical episode of *P. vivax*. Disentangling the contributions of relapses and reinfections is important to guide intervention selection, as certain interventions, such as LLINs, may be less effective during the short-term in settings where individuals frequently relapse.^18^

We projected that the distribution of LLINs in Bolívar would cause a greater proportional reduction in *P. falciparum* infections than *P. vivax* infections. The reduced impact on *P. vivax* occurred because most *P. vivax* infections were inferred to be relapses, not reinfections. Although LLINs can prevent onward transmission arising from relapses, they do not reduce the reservoir of hypnozoites acquired prior to intervention allocation. Therefore, short-term reductions in burden will be less for *P. vivax* than for *P. falciparum*, consistent with observations from a longitudinal study in Papua New Guinea.^18^

Relative to LLINs, MDA was projected to yield proportional reductions in incidence that were more comparable across *Plasmodium* spp. Increasing the duration of prophylaxis of the antimalarial drug provided a greater benefit for *P. falciparum* than *P. vivax*, because most *P. vivax* infections were caused by relapse, not reinfection. MDA that included an 8-aminoquinoline, such as PQ, is expected to cause a large reduction in *P. vivax* incidence by targeting the hypnozoite reservoir. Therefore, MDA that combines an antimalarial drug with a long duration of prophylaxis, such as dihydroartemisinin-piperaquine, and an 8-aminoquinoline, such as primaquine, to clear the hypnozoites may be an appropriate strategy to jointly address the burden of *P. falciparum* and *P. vivax*.

Our projected impact of MDA with PQ on the *P. vivax* burden in Bolívar revealed a tradeoff between the target coverage and the assumed level of PQ adherence. Low coverage scenarios with high PQ adherence were projected to have comparable levels of impact to high coverage scenarios with low PQ adherence, highlighting a potential role for focal MDA in Bolívar. With a lower target coverage, a focal MDA could ensure high rates of adherence among certain demographics at high risk of infection, such as gold miners, thereby addressing the large hypnozoite reservoir in the region.^6,8^ Success of focal MDA targeting individuals with occupational hazard of *Plasmodium* spp. infection was recently documented in a near-elimination setting in Costa Rica.^19^

The limited data on *Plasmodium* spp. prevalence and historical intervention use in the region could have affected our inferences and the resultant impact projections. Due to a lack of recent prevalence surveys in Bolívar, we calibrated our transmission models to weekly incidence data and geospatial *P. vivax* prevalence estimates from Battle and colleagues.^17^ Incidence data, however, is subject to issues of reporting and likely does not capture asymptomatic and low-density infections.^20^ The geospatial prevalence estimates are modelled quantities themselves and therefore must be interpreted with caution. Moreover, data on historical LLIN coverage was limited, which may have caused us to underestimate LLIN usage in the region and thus underestimate the time-varying reproduction numbers. Data on LLIN distribution during 2018-2019 was not available, though the increase in LLIN usage may help to explain the decline in transmission captured for *P. falciparum* in recent years. By potentially underestimating the reproduction numbers, we may also be overestimating the impact of interventions, so our projections should be considered as an upper bound of potential impact. Finally, we assumed that efficacy of CQ against *P. vivax* infection was high in Bolívar, so reduced efficacy on account of CQ resistance or poor adherence may reduce the impact of MDA. Nevertheless, we expect that the general relationships among intervention impact and target coverage, duration of prophylaxis, and assumed level of adherence should hold true.

Future intervention analyses in Bolívar would benefit from examining impact at finer spatial scales and accounting for ongoing demographic changes in this area. Transmission models calibrated at the municipality or parish levels could capture greater spatial heterogeneity in transmission risk^21^ and could be extended to realistically model the movement of hosts and parasites across administrative boundaries.^22^ Extending the framework developed in this analysis to accommodate finer spatial scales would continue efforts toward precision public health by providing policy makers with a model- and data-driven selection of targeted interventions to address the malaria epidemic in Venezuela.^23^

## Data Availability

Due to a data-sharing agreement, the underlying data can be publicly shared. All data requests should be made to LV.

## CONTRIBUTORS

JHH, LFC, LV, and TAP conceived of the study. JHH, ASS, and TAP performed the analyses. LFC and LV verified the data. JHH and TAP wrote the initial draft. All authors reviewed the manuscript and provided feedback.

## DECLARATION OF INTERESTS

TAP is a statistical reviewer for *The Lancet Infectious Diseases*. All other authors have no competing interests to declare.

## ACKNOWLEDGEMENTS

JHH acknowledges support from a National Science Graduate Research Fellowship and a Richard and Peggy Notebaert Premier Fellowship.

