## Supplementary Materials for "An epidemiological and intervention assessment of the malaria epidemic in Bolívar, Venezuela: a modelling study" (from another version of this preprint: version 1, 2022-04-21)

#### Table of Contents

Methods and Data ... 2
Data ... 2
Models ... 4
Inference Framework ... 18
Analyses ... 19
Sensitivity Analyses ... 20
Results ... 20
Daily Temperature Estimates in Bolívar ... 20
Simulation Study ... 21
Application to Epidemiological Data ... 24
Impact of Spatial Heterogeneity ... 28
Uncertainty in LLIN Action ... 31
Sensitivity of Impact Projections to Inference Replicates ... 32
References ... 34

### Methods and Data

#### Data

##### *Demographic Data*

To track changes in population over time (2000-2023), we used demographic projections generated by the United Nations World Population Prospects 2019, which included estimates of the total population size as well as the number of births, deaths, and net migration events for each year.[1] Demographic projections were available at the national level. To estimate the total population size and the number of births and deaths within Bolívar, Venezuela, we scaled the national-level data by the proportion of the population living in Bolívar during each year, which we computed from WorldPop population density estimates.[2] To interpolate changes in population on a weekly time scale, we divided the annual number of births, deaths, and net migration events by the number of weeks in each year. We then scaled each quantity to ensure that the net change in population obtained from this interpolation method matched the difference in the population of Bolívar from the start of one year to the start of the following year.

##### *Epidemiological Data*

*Incidence Data*

We used malaria incidence data reported by the Venezuelan Ministry of Health in Bolívar (Fig. S1). This data consisted of the weekly number of reported autochthonous cases of *Plasmodium falciparum* and *Plasmodium vivax* during 2000-2018, the weekly number of reported imported cases of *P. falciparum* and *P. vivax* during 2009-2018, and the weekly number of reported recrudescing and relapsing cases during 2008-2017. Each malaria case was diagnosed using light microscopy or a rapid diagnostic test. Case definitions under the Venezuelan Ministry of Health guidelines are provided in Table S1.

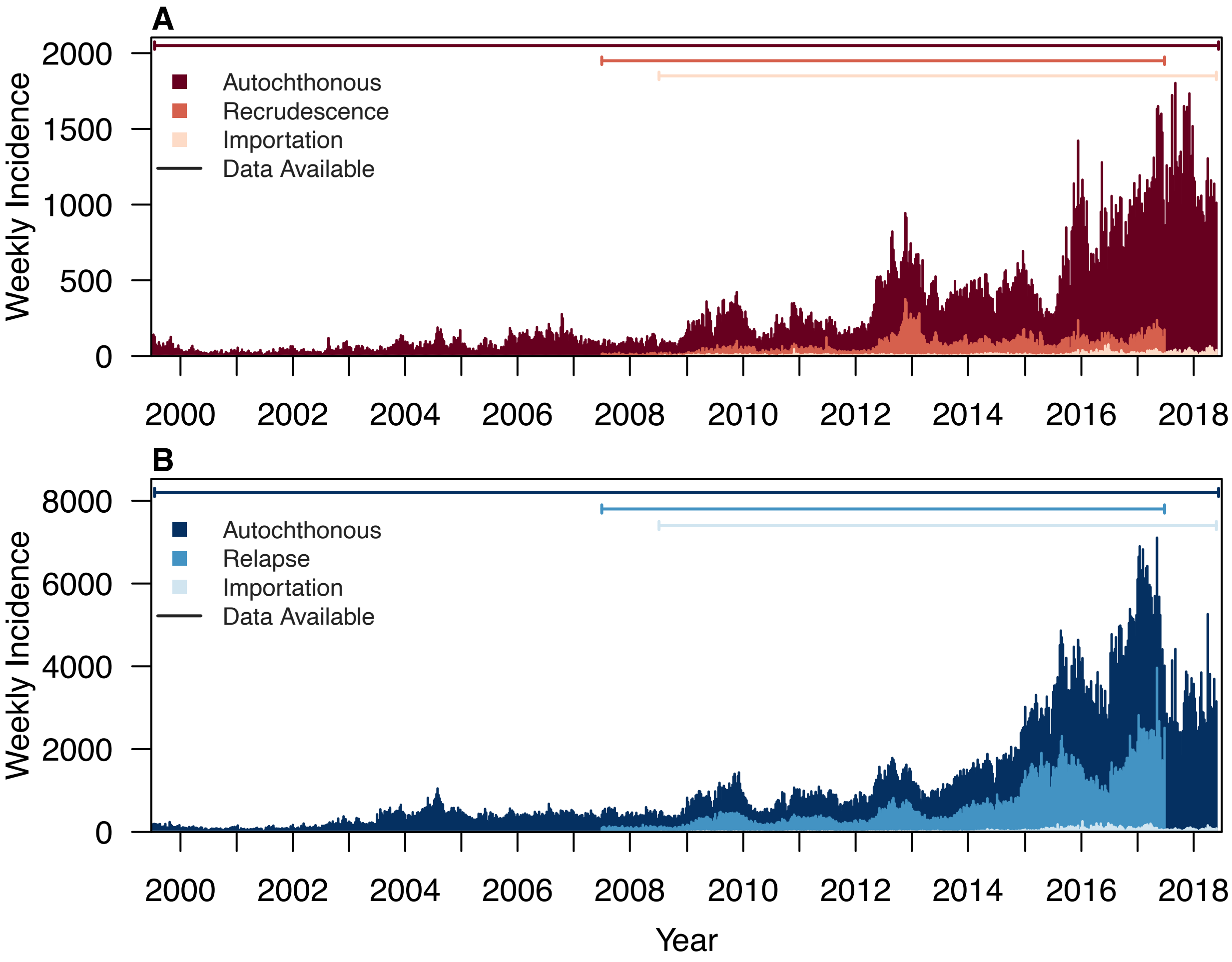

**Figure S1. Weekly reported malaria cases in Bolívar, Venezuela.** The weekly numbers of reported cases of (A) *P. falciparum* and (B) *P. vivax* are shown from 2000-2018. *P. falciparum* cases are classified as autochthonous, recrudescing, or imported. *P. vivax* cases are classified as autochthonous, relapsing, or imported. Each line denotes the time period over which weekly data for that case classification was available.

**Table S1. Venezuelan Ministry of Health Guidelines**

| Classification | Species | Definition |
|---|---|---|
| Autochthonous | *P. falciparum* or *P. vivax* | Infection was acquired within Bolívar, due to permanent or sustained malaria transmission |
| Importation | *P. falciparum* or *P. vivax* | Infection was acquired outside of Bolívar but was detected within Bolívar |
| Relapse | *P. vivax* | Infection presenting up to 90 days following radical cure of a primary infection |
| Recrudescence | *P. falciparum* | Infection presenting up to 30 days following the diagnosis of a primary infection |

Distinguishing between relapses, recrudescences, and reinfections of *P. vivax* is challenging.[3–5] The Venezuelan Ministry of Health classifies a *P. vivax* infection as a relapse if diagnosis occurred up to 90 days since cessation of radical cure therapy (i.e., 15 – 105 days following the initial diagnosis). Moreover, an infection is classified as a recrudescence if the diagnosis occurred within 30 days of the diagnosis of the primary infection. The weekly epidemiological data did not specify whether a recrudescence was caused by *P. falciparum* or *P. vivax* parasites. A previous study using time-to-event and genetic data estimated that recrudescences comprised 0·01% to 0·2% of all recurrent *P. vivax* infections following either treatment with high-dose primaquine and a blood-stage partner drug or treatment with chloroquine monotherapy, respectively.[5] Given this low rate of *P. vivax* recrudescence following treatment and the overlap in case definitions for relapses and recrudescences (Table S1), we assumed that all recrudescences were caused by *P. falciparum* parasites.

*Prevalence Data*

To account for underreporting of malaria infections, we used *P. vivax* prevalence estimates generated by the Malaria Atlas Project.[6,7] Battle *et al.*[6] estimated annual *P. vivax* prevalence for individuals 0-99 years of age during 2000-2017 at a 5km x 5km resolution. Annual prevalence estimates were also available for *P. falciparum* during the same time period.[8] However, the *P. falciparum* prevalence estimates from Weiss *et al.*[8] only considered individuals 2-10 years of age. Because the epidemic in Bolívar primary affects men aged 20-40 engaged in mining,[9,10] extrapolating the estimates from Weiss *et al.*[8] may have underestimated the prevalence in older age groups. Therefore, we chose to not incorporate *P. falciparum* prevalence estimates in our analysis.

We extracted *P. vivax* prevalence estimates for Bolívar, Venezuela using the *malariaAtlas* R package[7] and weighted them according to WorldPop population density estimates[2] to arrive at annual prevalence estimates for each municipality in Bolívar, Venezuela during 2000-2017 (Fig. S2).

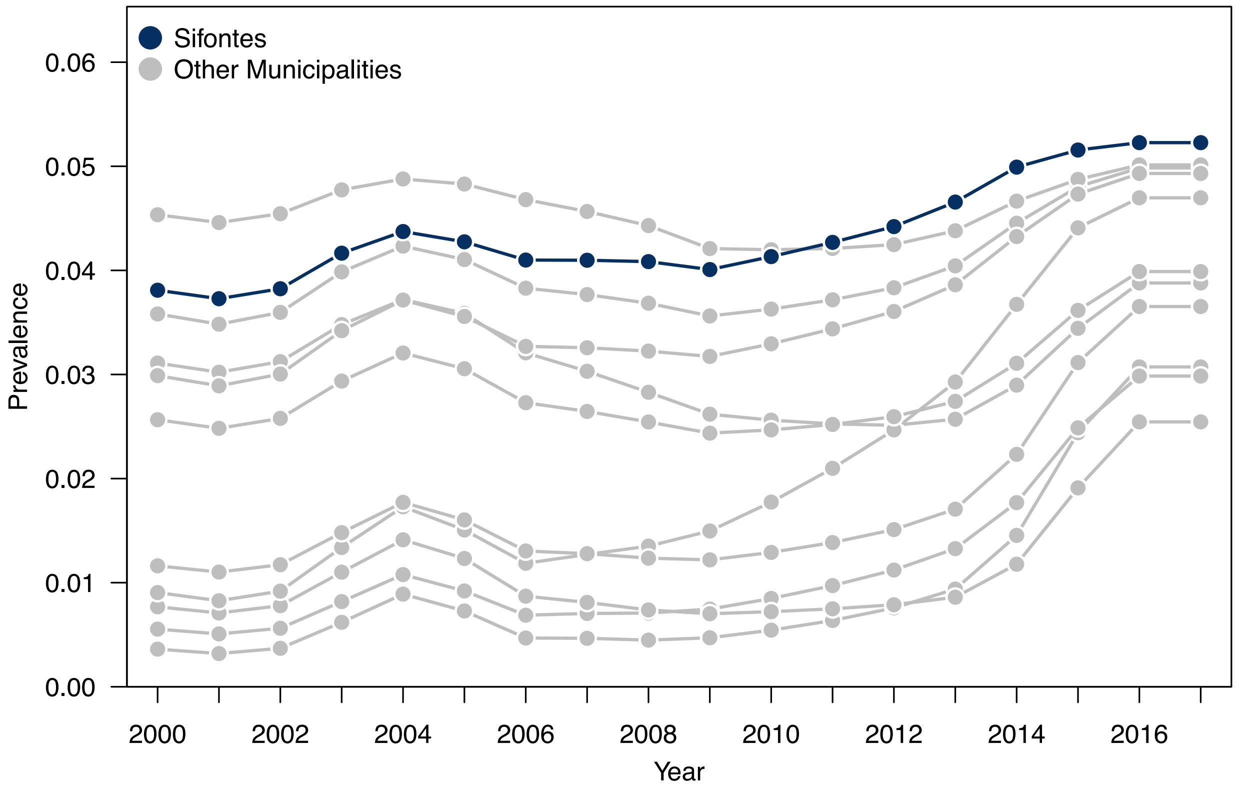


**Figure S2. Annual *P. vivax* prevalence estimates in Bolívar generated by the Malaria Atlas Project.**[6,7] Annual, population-weighted prevalence estimates are reported for all municipalities in Bolívar during 2000-2017. Sifontes municipality, the epicenter of the outbreak in Bolívar, is accented in blue.

***Temperature Data***

The transmission models for *P. vivax* and *P. falciparum* incorporated seasonality in transmission through an explicit dependence of vector and parasite traits on temperature. Therefore, the transmission models required daily temperature estimates for Bolívar. To inform these estimates, we acquired mean daily temperature readings during 1990-2019 from 157 weather stations in Venezuela, Colombia, Ecuador, Peru, Guyana, and Suriname from NOAA Climate Data Online.[11] The mean daily temperature readings informed a kriging model described below, which we used to estimate mean daily temperature in Bolívar.

***Entomological Data***

To validate entomological estimates derived from our transmission models, we used entomological data collected in Bolívar, Venezuela.[12,13] Moreno *et al.*[12] performed monthly landing catches between January 1999 and April 2000 in Sifontes, Bolívar and quantified the mean monthly number of *Anopheles darlingi* and *Anopheles marajoara* captured per person per night in the study area. Rubio-Palis *et al.*[13] performed mosquito catches using CDC light traps and mosquito magnets in the Lower Caura River Basin in Bolívar between March 2008 and January 2009 and reported the mean monthly number of *An. darlingi* and *An. nuneztovari* collected.

**Models**

***Temperature Model***

Mean daily temperature readings from weather stations were incomplete and required daily imputation. To address this shortcoming, we followed Siraj *et al.*[14] by using a kriging approach to estimate daily temperature surfaces and weighting those surfaces according to gridded population density estimates to obtain mean daily temperature estimates for Bolívar.

To inform our kriging approach, we used the Global 30 Arc-Second Elevation (GTOPO30) and the WorldClim long-term average temperature surfaces as covariates.[15,16] We then fitted a kriging model with these covariates to estimate a mean daily temperature surface for each day between January 1st, 1995 and October 3rd, 2019. This model was represented as

$$T(x) = \mu(x) + S(x) + \varepsilon, \quad \text{(S1)}$$

where $T(x)$ is the temperature at location $x$, $\mu(x)$ is the fixed effect at location $x$, $S(x)$ is a stationary Gaussian process that is described by a variance and correlation function, and ε is the error term.[14] Following Siraj *et al.*,[14] we considered the temperature on each day to be independent from the temperatures on all other days and fitted the model in eq. (S1) to the corresponding temperature readings from that day. By doing so, we were able to estimate the time-series of mean daily temperature surfaces in Bolívar. Finally, we aggregated the mean daily temperature estimates to the municipality and state levels by weighting the predicted surfaces according to gridded population density estimates.[2] At time points for which no temperature data were available, we imputed the mean daily temperature as the 31-day moving average. For temperature in future years (i.e., 2020-2023), we assumed that there was no change in the long-term trend in temperature and set the daily temperature for each day equal to the average temperature on that day over the last three years.

***Transmission Models***

To model transmission of *P. falciparum* and *P. vivax* on a weekly timescale in Bolívar, Venezuela, we adapted a renewal equation model with features appropriate for this epidemiological setting.[17] Alternative versions of this model have been successfully applied in the context of measles,[18] cholera,[19] and chikungunya,[20] among other instances. One study previously developed a similar model to describe *P. falciparum* transmission in Western Kenya.[21] However, we did not make use of this model and instead developed our own model for two reasons. First, the existing model was not structured to model *P. vivax* transmission and the relapses therein. Second, the model did not account for recrudescences that may arise from *P. falciparum* treatment failure and that were documented in the incidence data from Bolívar. Accordingly, transmission models with features tailored to the epidemiological dynamics in Bolívar were needed.

*P. falciparum Transmission Model*

Under our model, the total number of new autochthonous infections of *P. falciparum* during week $t$ is

$$I_t^{(total)} = \sum_{\tau=1}^{t-1} \boldsymbol{R}(t|t-\tau)\frac{I'_{t-\tau}}{N_t}S_t\,. \quad \text{(S2)}$$

In eq. (S2), $\boldsymbol{R}(t|t-\tau)$ is the reproduction number exerted by individuals infected during week $t-\tau$ on susceptible individuals during week $t$, $I'_{t-\tau}$ is the total number of new infections during week $t-\tau$, $N_t$ is the population size of Bolívar during week $t$, and $S_t$ is the number of susceptible individuals at the start of week $t$. We calculated $I'_{t-\tau}$ in eq. (S2) as

$$I'_{t-\tau} = I_{t-\tau}^{(total)} + \xi_{t-\tau}^{(total)} + \iota_{t-\tau}, \quad \text{(S3)}$$

where $\xi_{t-\tau}^{(total)}$ and $\iota_{t-\tau}$ are the respective number of new recrudescing and imported infections during week $t-\tau$. In eq. (S3), the number of new recrudescing infections is a state variable in our model, while the number of new imported infections was informed by data from the Venezuelan Ministry of Health. The number of new autochthonous infections, $I_t^{(total)}$, can be sub-divided into those that ultimately present at a health clinic, $I_t^{(rep)}$, and those that do not, $I_t^{(unrep)}$. We calculated the former as $\rho(t)I_t^{(total)}$ and the latter as $\left(1-\rho(t)\right)I_t^{(total)}$, where $\rho(t)$ is the probability of seeking treatment at a health clinic conditional on infection occurring on week $t$ and was modeled as a cubic spline with two knots. The values of these knots were estimated during model fitting.

The number of new autochthonous infections beginning antimalarial treatment in a health clinic during week $t$ is calculated as

$$T_t^{(I)} = \sum_{\tau=1}^{t-1} IDP(\tau)I_{t-\tau}^{(rep)}, \quad \text{(S4)}$$

where $IDP(\tau)$ is the probability that the duration from infection-to-detection is τ weeks and $I_{t-\tau}^{(rep)}$ is the number of new autochthonous infections acquired on week $t-\tau$ that ultimately present at a health clinic. We further assumed that a proportion $\varrho(t)$ of individuals not seeking treatment in the health clinic received medication through other means, where $\chi(t)$ was estimated as a scalar multiple of $\rho(t)$. Therefore, the number of new autochthonous infections that self-medicated during week *t* is calculated as

$$I_t^{(unrep,T)} = \sum_{\tau=1}^{t-1} FDP(\tau)\chi(t-\tau)I_{t-\tau}^{(unrep)}. \quad \text{(S5)}$$

We modeled non-adherence to antimalarial treatment by assuming that a proportion $\theta_{ACT}$ of treated individuals completely adhere to antimalarial treatment. We assumed that non-adherence to antimalarial treatment resulted in a recrudescing infection and modeled the total number of new recrudescing infections during week *t* with a reported primary infection as

$$\xi_t^{(total,1^\circ\, rep)} = (1-\theta_{ACT})T_{t-\gamma_\xi}^{(I)}\mathbb{I}(t > \gamma_\xi), \quad \text{(S6)}$$

where $\gamma_\xi$ is the time from treatment to the onset of a recrudescing infection and was assumed to be 2 weeks,[22] and $\mathbb{I}(t > \gamma_\xi)$ is an indicator function equal to one if $t > 2$ and zero otherwise. Similarly, the total number of new recrudescing infections during week *t* without a reported primary infection was calculated as

$$\xi_t^{(total,1^\circ\, unrep)} = (1-\theta_{ACT})I_{t-\gamma_\xi}^{(unrep,T)}\mathbb{I}\big(t > \gamma_\xi\big). \quad \text{(S7)}$$

As with new autochthonous infections, we assumed that the number of new recrudescing infections that ultimately present at a health clinic, $\xi_t^{(rep)}$, depended upon $\rho(t)$ and was equal to $\rho(t)\xi_t^{(total)}$. Similarly, the number of new recrudescing infections that did not present at a health clinic, $\xi_t^{(unrep)}$, was $\big(1-\rho(t)\big)\xi_t^{(total)}$. This calculation applied to recrudescences arising from both reported and unreported primary infections.

In the absence of molecular diagnosis, distinguishing between recrudescences and re-infections is challenging. According to Venezuelan Ministry of Health guidelines, a *P. falciparum* infection is classified as recrudescing if clinical presentation occurred within 30 days of a prior diagnosis (Table S1). To make our modeled quantities comparable to the Bolívar data, we modeled recrudescing infections consistent with this definition. Specifically, we calculated the number of new and correctly classified recrudescing infections that appeared in the health clinic on week *t* as

$$T_t^{(\xi,c)} = \sum_{\tau=1}^{\min\{2,t-1\}} FDP(\tau)\,\xi_{t-\tau}^{(rep,1^\circ\, rep)}, \quad \text{(S8)}$$

where $FDP(\tau)$ is the probability that time from fever onset to clinical presentation was τ weeks. Because we assumed that the onset of fever coincided with the onset of recrudescing infection, the time from fever onset to clinical presentation could not exceed two weeks for the case to be correctly classified as a recrudescing infection according to Venezuelan Ministry of Health guidelines. Thus, any recrudescing infection that presented outside this time window, as well as any recrudescing infection with an unreported primary infection, was misclassified as a new infection, and we calculated this quantity as

$$T_t^{(\xi,m)} = \sum_{\tau=3}^{t-1} FDP(\tau)\,\xi_{t-\tau}^{(rep,1^\circ rep)} + \sum_{\tau=1}^{t-1} FDP(\tau)\xi_{t-\tau}^{(rep,1^\circ\, unrep)}. \quad \text{(S9)}$$

Eq. (S2-S9) describe the transition of individuals through the infected and recrudescing compartments along with their detection by the health system. The rate at which individuals recovered from *P. falciparum* infection depended upon on antimalarial drug treatment. We modeled the number of newly recovered individuals as

$$R_t = \sum_{\tau=1}^{t-1} \gamma_U(\tau) \left( I_{t-\tau}^{(unrep,U)} + \xi_{t-\tau}^{(unrep)} \right) + \sum_{\tau=1}^{t-1} \gamma_T(\tau) \left( \theta_{ACT} (T_{t-\tau}^{(I)} + I_{t-\tau}^{(unrep,T)}) + T_{t-\tau}^{(\xi,c)} + T_{t-\tau}^{(\xi,m)} \right). \quad \text{(S10)}$$

In eq. (S10), the first summation quantifies the number of individuals who did not present at a health clinic and cleared their infections naturally during week *t*. The second summation quantifies the number of individuals who cleared their infections during week *t* as a result of antimalarial drug treatment.

We calculated the number of susceptible individuals at the start of week *t* as

$$S_t = S_{t-1} + B_{t-1} - D_{t-1} \frac{S_{t-1}}{N_{t-1}} - I_{t-1}^{(total)} + \sum_{\tau=1}^{t-1} \kappa_\tau R_{t-\tau}, \quad \text{(S11)}$$

where $B_{t-1}$ and $D_{t-1}$ are the number of births and deaths that occurred during week *t* – 1. It is important to note that we considered only the fraction of deaths among susceptible individuals (i.e., $S_{t-1}/N_{t-1}$) and therefore did not model deaths due to malaria. We assumed that recovered individuals experience an exponentially distributed duration of immunity that temporarily protects them from re-infection. To account for this, the summation involving $\kappa_\tau$ in eq. (S11) denotes the number of recovered individuals who are susceptible at the start of week *t*. The parameter $\kappa_\tau$ is defined as the proportion of individuals who lost their immunity $\tau$ weeks after clearing their infection and is calculated as

$$\kappa_\tau = 1 - e^{-\lambda\tau}. \quad \text{(S12)}$$

In eq. (S12), $\lambda^{-1}$ is the mean duration of immunity following clearance of infection and is assumed to be one week (Table S3).

**Table S2. State variables for the *P. falciparum* transmission model.**

| State Variable | Description |
|---|---|
| $S_t$ | Number of susceptible individuals at the start of week *t* |
| $B_t$ | Number of births that occurred during week *t* |
| $D_t$ | Number of deaths that occurred during week *t* |
| $N_t$ | Size of population at the start of week *t* |
| $I_t^{(rep)}$ | Number of new autochthonous infections during week *t* that ultimately present at a health clinic |
| $I_t^{(unrep)}$ | Number of new autochthonous infections during week *t* that do not present at a health clinic |
| $I_t^{(unrep,T)}$ | Number of new autochthonous infections during week *t* that do not present at a health clinic but self-medicate |
| $I_t^{(unrep,U)}$ | Number of new autochthonous infections during week *t* that do not present at a health clinic and do not self-medicate |
| $T_t^{(I)}$ | Number of new autochthonous infections receiving treatment during week *t* |
| $\xi_t^{(rep,1^\circ rep)}$ | Number of new recrudescing infections during week *t* with a reported primary infection that ultimately present at a health clinic |
| $\xi_t^{(rep,1^\circ unrep)}$ | Number of new recrudescing infections during week *t* with an unreported primary infection that ultimately present at a health clinic |
| $\xi_t^{(unrep,1^\circ rep)}$ | Number of new recrudescing infections during week *t* with a reported primary infection that do not present at a health clinic |
| $\xi_t^{(unrep,1^\circ unrep)}$ | Number of new recrudescing infections during week *t* with an unreported primary infection that do present at a health clinic |
| $T_t^{(\xi,c)}$ | Number of new recrudescing infections receiving treatment during week *t* that are correctly classified as recrudescing infections |
| $T_t^{(\xi,m)}$ | Number of new recrudescing infections receiving treatment during week *t* that are incorrectly classified as autochthonous infections |
| $\iota_t$ | Number of new imported infections during week *t* |
| $R_t$ | Number of new recovered individuals during week *t* |

**Table S3. Parameters of the *P. falciparum* transmission model.**

| Parameter | Description | Value | Source |
|---|---|---|---|
| $\Lambda(t)$ | Force of infection at time *t* | Estimated | |
| $\rho(t)$ | Probability of seeking treatment conditional on infection on time *t* | Estimated | |
| $\varrho(\tau)$ | Probability of self-medicating conditional on infection on time *t* | Estimated | |
| $\gamma_U$ | Duration of untreated *P. falciparum* infection | Empirical distribution (Mean: 24·5 weeks) | [23] |
| $\gamma_T$ | Duration of treated *P. falciparum* infection | Empirical distribution (Mean: 4·6 weeks) | [24] |
| $\gamma_\xi$ | Time from treatment to onset of recrudescing infection | 2 weeks | [22] |
| $IDP$ | Duration of time from infection-to-detection | Empirical distribution (Mean: 2·8 weeks) | [25] |
| $FDP$ | Duration of time from fever to detection | Empirical distribution (Mean: 1 week) | [25] |
| $\theta_{ACT}$ | Proportion of individuals adhering to ACT treatment | Estimated | |
| $\lambda^{-1}$ | Mean duration of immunity | 1 week | Assumed |

*Plasmodium vivax Transmission Model*

To model *P. vivax* transmission over time, we used a model that was similar to the one that we used for *P. falciparum*. The main difference between these models is that we account for the dormant, liver-stage parasites of *P. vivax* known as hypnozoites, which cause relapsing infections. A full list of the state variables along with the model parameters for the *P. vivax* transmission model are provided in Tables S4 and S5.

Under this *P. vivax* transmission model, the number of new autochthonous infections on week *t* is calculated as

$$I_{L,t}^{(total)} = \sum_{\tau=1}^{t-1}\left(R^{(T)}(t|t-\tau)\frac{I_{t-\tau}^{T'}}{N_t} + R^{(U)}(t|t-\tau)\frac{I_{t-\tau}^{U'}}{N_t}\right)\left(S_{0,t} + S_{L,t}^{(1^\circ\, unrep)} + S_{L,t}^{(1^\circ\, rep)}\right). \quad \text{(S13)}$$

In eq. (S13), $R^{(T)}(t|t-\tau)$ and $R^{(U)}(t|t-\tau)$ are the respective reproduction numbers exerted by treated and untreated individuals infected during week $t$ - $\tau$ on susceptible individuals infected during week *t*, $I_{t-\tau}^{T'}$ and $I_{t-\tau}^{U'}$ are the respective total number of treated and untreated individuals that were infected during week $t$ - $\tau$, and $N_t$ is the total population size during week *t*. Moreover, $S_{0,t}$ is the number of susceptible individuals at the start of week *t* that do not have liver-stage parasites, $S_{L,t}^{(1^\circ\, unrep)}$ is the number of susceptible individuals at the start of week *t* that have liver-stage parasites attributable to an unreported primary infection, and $S_{L,t}^{(1^\circ\, rep)}$ is the number of susceptible individuals at the start of week *t* that have liver-stage parasites attributable to a reported primary infection. We calculated $I_{t-\tau}^{T'}$ in eq. (S13) as

$$I_{t-\tau}^{T'} = I_{t-\tau}^{(rep)} + \chi(t-\tau)I_{t-\tau}^{(unrep)} + \xi_{t-\tau}^{(rep)} + \chi(t-\tau)\xi_{t-\tau}^{(unrep)} + \iota_{t-\tau}, \quad (S14)$$

where $I_{t-\tau}^{(rep)}$ and $\xi_{t-\tau}^{(rep)}$ are the respective number of new autochthonous infections and relapses that occurred during week $t-\tau$ and that will present in the health clinic. By contrast, $I_{t-\tau}^{(unrep)}$ and $\xi_{t-\tau}^{(unrep)}$ are the respective number of new autochthonous infections and relapses that occurred during $t-\tau$ and that will not present in the health clinic, and $\chi(t-\tau)$ is the proportion of these unreported infections and relapses on week *t* that will self-medicate. We similarly calculated $I_{t-\tau}^{U'}$ in eq. (S13) as

$$I_{t-\tau}^{U'} = (1-\chi(t-\tau))\left(I_{t-\tau}^{(unrep)} + \xi_{t-\tau}^{(unrep)}\right). \quad \text{(S15)}$$

We calculated the number of reported and unreported autochthonous infections on the basis of whether each individual was likely to experience symptoms and, if experiencing symptoms, whether each individual was likely to seek treatment at a health clinic. We defined $\phi_{D,Max}$ as the maximum probability that an individual would experience symptoms and applied this quantity to individuals who had no recent *P. vivax* infection (i.e., $S_{0,t}$), and we defined $\phi_{D,Min}$ as the minimum probability that an individual would experience symptoms and applied this quantity to individuals with recent *P. vivax* infection (i.e., $S_{L,t}^{(1^\circ\, unrep)}$ and $S_{L,t}^{(1^\circ\, rep)}$). Furthermore, we defined $\rho(t)$ as the probability that a symptomatic individual infected during week *t* would seek treatment in the health clinic. We further distinguished newly infected individuals that ultimately show up in the health system on the basis of whether they had a previously detected primary infection, $I_{L,t}^{(rep;\, 1^\circ\, rep)}$, or did not have a previously detected primary infection, $I_{L,t}^{(rep;\, 1^\circ\, unrep)}$. By doing so, we were able to address potential errors in classification of cases as autochthonous or relapse. The number of new, correctly classified autochthonous cases beginning chloroquine treatment along with effective radical cure therapy is

$$T_{0,t}^{(c)} = \theta_{PQ}\varphi_{PQ}\sum_{\tau=1}^{t-1} IDP(\tau)\left(\theta_{C_I} I_{L,t-\tau}^{(rep;1^\circ\, rep)} + I_{L,t-\tau}^{(rep;1^\circ\, unrep)}\right), \quad \text{(S16)}$$

where $\theta_{PQ}$ is the proportion of cases receiving radical cure therapy (i.e., primaquine), $\varphi_{PQ}$ is the assumed efficacy of primaquine, $IDP(\tau)$ is the probability that the duration from infection-to-detection is $\tau$ weeks, and $\theta_{C_I}$ is the proportion of autochthonous infections with a previously detected primary infection that are correctly classified. The latter quantity is calculated as proportion of autochthonous infections with a previously detected primary infection that are expected to present outside of the 90-day window for classification of cases as relapses as defined in Table S1 and depends upon the rate at which individuals in the population are reinfected and the infection-to-detection period. The number of new autochthonous cases beginning chloroquine treatment along with effective radical cure that are incorrectly classified as relapses is

$$T_{0,t}^{(m)} = \theta_{PQ}\varphi_{PQ}\left(1-\theta_{C_I}\right)\sum_{\tau=1}^{t-1} IDP(\tau) I_{L,t-\tau}^{(rep;1^\circ\, rep)}. \quad \text{(S17)}$$

Infected individuals who presented at a health clinic either did not receive radical cure or did not fully adhere to radical cure were assumed to be latently infected with hypnozoites and were thus able to relapse. We calculated the number of correctly classified autochthonous cases with liver-stage parasites as

$$T_{L,t}^{(c)} = \left(1-\theta_{PQ}\varphi_{PQ}\right)\sum_{\tau=1}^{t-1} IDP(\tau)\left(\theta_{C_I} I_{t-\tau}^{(rep;1^\circ\, rep)} + I_{t-\tau}^{(rep;1^\circ\, unrep)}\right) \quad \text{(S18)}$$

and the number of new autochthonous cases with liver-stage parasites that were misclassified as relapses as

$$T_{0,t}^{(m)} = \left(1-\theta_{PQ}\varphi_{PQ}\right)\left(1-\theta_{C_I}\right)\sum_{\tau=1}^{t-1} IDP(\tau) I_{L,t-\tau}^{(rep;1^\circ\, rep)}. \quad \text{(S19)}$$

In eqs. (S18) and (S19), $1-\theta_{PQ}\varphi_{PQ}$ is the probability of not receiving effective radical cure (i.e., the probability of having liver-stage parasite after presenting at a health clinic). Following clearance of the bloodstream infection, treated individuals ($T_{0,t}^{(c)}$ and $T_{0,t}^{(m)}$) receiving effective radical cure enter into the recovered class *R* where they experience a period of temporary immunity. Following clearance of their bloodstream infection, unreported infected individuals enter into $S_{L,t}^{(1^\circ\, unrep)}$, and treated individuals with liver-stage parasites ($T_{L,t}^{(c)}$ and $T_{L,t}^{(m)}$) enter into $S_{L,t}^{(1^\circ\, rep)}$, which tracks the number of individuals who cleared their reported primary bloodstream infection and can relapse.

Following White *et al.*,[26,27] we assumed that individuals relapse with rate $\gamma_\xi$. Therefore, the number of unreported relapses that occurred on week *t* was calculated as

$$\xi_t^{(unrep)} = \left(1-e^{-\gamma_\xi}\right)\left(\left(1-\phi_{D,Min}\rho(t)\right)S_{L,t}^{(1^\circ\, unrep)} + \left(1-\phi_{D,Max}\rho(t)\right)S_{L,t}^{(1^\circ\, rep)}\right). \quad \text{(S20)}$$

In eq. (S20), the first term in the product is the probability of relapsing during week *t*, and the second term in the product is the number of susceptible individuals that would not have a reported relapse during week *t*. Similar to eq. (S20), we calculated the number of new relapses occurring on week *t* that would present at a health clinic as

$$\xi_{1^\circ\, unrep,t}^{(rep)} = \phi_{D,Min}\rho(t)\left(1-e^{-\gamma_\xi}\right)S_{L,t}^{(1^\circ\, unrep)} \quad \text{(S21)}$$

$$\xi_{1^\circ\, rep,t}^{(rep)} = \phi_{D,Max}\rho(t)\left(1-e^{-\gamma_\xi}\right)S_{L,t}^{(1^\circ\, rep)}. \quad \text{(S22)}$$

Unlike for unreported relapses, we distinguished between reported relapses that arose from a reported versus unreported primary infection. We made this distinction to be consistent with the case definition for a relapse that stated that the primary infection must be reported (Table S1). Therefore, the number of relapses that were correctly classified as relapses on week *t* was

$$T_{\xi,t}^{(c)} = \sum_{\tau=1}^{t-1} \theta_{C_\xi} FDP(\tau)\xi_{1^\circ\, rep,t-\tau}^{(rep)}, \quad \text{(S23)}$$

where $\theta_{C_\xi}$ is the proportion of relapses that are correctly classified as relapses and $FDP(\tau)$ is the probability that the duration of time from fever-onset to detection is τ weeks. By comparison, the number of relapses that were misclassified as autochthonous infections during week *t* was

$$T_{\xi,t}^{(m)} = \sum_{\tau=1}^{t-1} FDP(\tau)\left(\left(1-\theta_{C_\xi}\right)\xi_{1^\circ\, rep,t-\tau}^{(rep)} + \xi_{1^\circ\, unrep,t-\tau}^{(rep)}\right). \quad \text{(S24)}$$

Consistent with Ministry of Health guidelines (Table S1), we assumed in eq. (24) that all relapsing individuals that presented to a health clinic and had an unreported primary infection were misclassified as an autochthonous infection.

Eqs. (S13)-(S24) describe the transition of individuals into the infected and relapsing compartments and determine whether or not they appear in the health system. The rate at which individuals recover from their infections depended upon whether or not they received antimalarial treatment. To enter into the recovered compartment *R*, an individual must have cleared both their primary bloodstream infection and their latent infection with hypnozoites. The number of newly recovered individuals during week *t* was thus modeled as

$$R_t = (1-e^{-\gamma_L})\left(S_{L,t}^{(1^\circ\, rep)} + S_{L,t}^{(1^\circ\, unrep)}\right) + \sum_{\tau=1}^{t-1}\gamma_T(\tau)\left(T_{0,t-\tau}^{(c)} + T_{0,t-\tau}^{(m)} + \theta_{PQ}\varphi_{PQ}\left(T_{\xi,t-\tau}^{(c)} + T_{\xi,t-\tau}^{(m)}\right)\right). \quad \text{(S25)}$$

In eq. (S25), the first term in the summation is the number of susceptible individuals that clear their hypnozoites naturally through the death of the hypnozoite or the infected hepatocyte, where $\gamma_L$ is the rate of clearance of hypnozoites. The second term in eq. (S25) is the number of treated individuals who cleared both their primary bloodstream infection and liver-stage parasites during week *t*, and the $\gamma_T(\tau)$ is the probability that an individual clears their bloodstream infection τ weeks after beginning treatment.

We then modeled the number of susceptible individuals $S_{L,t}^{(1^\circ\, unrep)}$ at the start of week *t* that had a previously unreported primary infection as

$$\begin{aligned}
S_{L,t}^{(1^\circ\, unrep)} = {} & S_{L,t-1}^{(1^\circ\, unrep)} + \sum_{\tau=1}^{t-2}\gamma_U(\tau)(1-\chi(t-1))\left(I_{L,t-1-\tau}^{(unrep)} + \xi_{t-1-\tau}^{(unrep)}\right) \\
& + \sum_{\tau=1}^{t-2}\gamma_T(\tau)\left(\left(1-\theta_{PQ}\varphi_{PQ}\right)\left(T_{\xi,t-1-\tau}^{(c)} + T_{\xi,t-1-\tau}^{(m)}\right) + \varrho(t-1)\left(I_{L,t-1-\tau}^{(unrep)} + \xi_{t-1-\tau}^{(unrep)}\right)\right) \\
& - I_{L,t-1}^{(total)}\left(\frac{S_{L,t-1}^{(1^\circ\, unrep)}}{S_{L,t-1}^{(1^\circ\, unrep)} + S_{L,t-1}^{(1^\circ\, rep)} + S_{0,t-1}}\right) - (1-e^{-\gamma_\xi})S_{L,t-1}^{(1^\circ\, unrep)} - (1-e^{-\gamma_L})S_{L,t-1}^{(1^\circ\, unrep)} \\
& - D_{t-1}\left(\frac{S_{L,t-1}^{(1^\circ\, unrep)}}{N_{t-1}}\right). \quad \text{(S26)}
\end{aligned}$$

In eq. (S26), the second term is the number of unreported autochthonous and relapsing infections that cleared their infections naturally by the start of the week *t*, and the third term is the number of the treated relapsing infections that cleared their primary bloodstream infection but did not cleared their hypnozoites by the start of week *t*. The remaining terms in eq. (S26) represent the loss of individuals due to autochthonous infection (4$^{th}$ term), relapsing infection (5$^{th}$ term), natural clearance of the hypnozoites (6$^{th}$), and death (7$^{th}$ term). We similarly calculated the total number of susceptible individuals at the start of week *t* that had a previously reported primary infection as

$$\begin{aligned}
S_{L,t}^{(1^\circ\, rep)} = {} & S_{L,t-1}^{(1^\circ\, rep)} + \sum_{\tau=1}^{t-1}\gamma_T(\tau)\left(T_{L,t-\tau}^{(c)} + T_{L,t-\tau}^{(m)}\right) - I_{L,t-1}^{(total)}\left(\frac{S_{L,t-1}^{(1^\circ\, rep)}}{S_{L,t-1}^{(1^\circ\, unrep)} + S_{L,t-1}^{(1^\circ\, rep)} + S_{0,t-1}}\right) - (1-e^{-\gamma_\xi})S_{L,t-1}^{(1^\circ\, rep)} \\
& - (1-e^{-\gamma_L})S_{L,t-1}^{(1^\circ\, rep)} - D_{t-1}\left(\frac{S_{L,t-1}^{(1^\circ\, rep)}}{N_{t-1}}\right). \quad \text{(S27)}
\end{aligned}$$

In eq. (S27), the second term is the number of treated individuals that clear their bloodstream infection. The remaining terms represent the loss of individuals due to autochthonous infection (3rd term), relapsing infection (4th term), natural clearance of the hypnozoites (5th term), and death (6th term). Finally, we calculated the number of susceptible individuals without hypnozoites at the start of week *t* as

$$S_{0,t} = S_{0,t-1} + B_{t-1} + \sum_{\tau=0}^{t-1} \kappa_\tau R_{t-\tau} - I_{L,t-1}^{(total)} \left( \frac{S_{0,t-1}}{S_{L,t-1}^{(1^\circ\, unrep)} + S_{L,t-1}^{(1^\circ\, rep)} + S_{0,t-1}} \right) - D_{t-1} \left( \frac{S_{0,t-1}}{N_{t-1}} \right). \quad \text{(S28)}$$

In eq. (S28), $B_{t-1}$ is the number of births in week *t* – 1, and the second term is the number of recovered individuals that return to the susceptible compartment at the start of week *t*. A description of the process that governs the loss of immunity is provided in eq. (S12).

**Table S4. State variables for *P. vivax* transmission model**

| State Variable | Description |
|---|---|
| $S_{0,t}$ | Susceptible individuals at the start of week *t* without liver-stage parasites |
| $B_t$ | Births that occurred during week *t* |
| $D_t$ | Deaths that occurred during week *t* |
| $N_t$ | Population at the start of week *t* |
| $I_{L,t}^{(rep;1^\circ\, rep)}$ | New infections during week *t* with a previously reported primary infection that will show up in a health clinic |
| $I_{L,t}^{(rep;1^\circ\, unrep)}$ | New infections during week *t* with a previously unreported primary infection that will show up in a health clinic |
| $I_{L,t}^{(unrep)}$ | New infections during week *t* that will not show up in a health clinic |
| $T_{0,t}^{(c)}$ | New infections receiving treatment during week *t* without hypnozoites that are correctly classified as autochthonous infections |
| $T_{0,t}^{(m)}$ | New infections receiving treatment during week *t* without hypnozoites that are misclassified as relapsing infections |
| $T_{L,t}^{(c)}$ | New infections receiving treatment during week *t* with hypnozoites that are correctly classified as autochthonous infections |
| $T_{L,t}^{(m)}$ | New infections receiving treatment during week *t* with hypnozoites that are misclassified as relapsing |
| $\xi_t^{(unrep)}$ | New relapses during week *t* that do not present at a health clinic |
| $\xi_{1^\circ\, rep,t}^{(rep)}$ | New relapses during week *t* with reported primary infection that will present at a health clinic |
| $\xi_{1^\circ\, unrep,t}^{(rep)}$ | New relapses during week *t* with unreported primary infection that will present at a health clinic |
| $T_{\xi,t}^{(c)}$ | New relapses receiving treatment during week *t* that are correctly classified as relapsing infections |
| $T_{\xi,t}^{(m)}$ | New relapses receiving treatment during week *t* that are misclassified as autochthonous infections |
| $S_{L,t}^{(1^\circ\, rep)}$ | Susceptible individuals with liver-stage parasites at start of week *t* that had a previously reported primary infection |
| $S_{L,t}^{(1^\circ\, unrep)}$ | Susceptible individuals with liver-stage parasites at start of week *t* that had a previously unreported primary infection |

**Table S5. Parameters of the *P. vivax* transmission model**

| Parameter | Description | Value | Source |
|---|---|---|---|
| $\chi(t)$ | Proportion of unreported infections on week *t* that will self-medicate | Estimated | |
| $\theta_{PQ}$ | Proportion of cases receiving PQ treatment | Estimated | |
| $\varphi_{PQ}$ | Efficacy of PQ treatment | 55·3% | 27 |
| $\gamma_\xi$ | Rate of relapse | 0·132 wk$^{-1}$ | 28 |
| $\gamma_L$ | Rate of clearance of hypnozoites | 0.03 wk$^{-1}$ | 27 |
| $\theta_{C_I}$ | Proportion of correctly classified autochthonous infections | Calculated | |
| $\theta_{C_\xi}$ | Proportion of correctly classified relapses | Calculated | |
| $\gamma_T$ | Duration of treated *P. vivax* infection | 1 wk | 27 |

*Computing the Reproduction Number*

To compute the time-varying reproduction number, we estimated the expected number, $\boldsymbol{R}(t_l|t_i)$, of secondary infections occurring on day $t_l$ arising from each primary infection that occurred on day $t_i$. This approach generalizes the Ross-MacDonald formula for the reproduction number and explicitly accounted for temporal variation in each component of the transmission cycle.[29] By adopting this mechanistic approach, we were able to consider the effects of seasonal fluctuations in mosquito density and temperature variation on transmission and could more directly project the impact of interventions by considering the effects of the interventions on the individual components of the transmission cycle.

In the following sections, we present the methodology to compute the time-varying reproduction number for *P. falciparum*. Due to the lack of a detailed within-host, mechanistic model for *P. vivax*, computing an equivalent time-varying reproduction number tailored to the biology of *P. vivax* was challenging. As such, we computed the time-varying reproduction number for *P. vivax* as a scalar multiple of the time-varying reproduction number for *P. falciparum*, with that scalar multiple estimated as a parameter during model-fitting.

*Human-To-Mosquito Transmission*

The first component of the transmission cycle involves transmission of gametocytes from a primary infection to blood-feeding mosquitoes. Therefore, we needed to calculate the expected number, $n_{HM}(t_j|t_i)$, of blood-feeding mosquitoes that were infected on day $t_j$ from primary infection that began on $t_i$.

To generate this time-varying quantity, we modeled the time-course of a *P. falciparum* infection following inoculation of a primary individual with sporozoites on day $t_i$. Consistent with Huber *et al.*,[25] we assumed that a period of 6 days must elapse for asexual parasites to emerge from the liver[30] and that, during this period prior to emergence from the liver, no transmission could occur. Upon emergence of asexual parasites from the liver, a primary bloodstream infection occurs, and an infected individual can transmit gametocytes to blood-feeding mosquitoes. To model changes in the probability of transmission over time, we used a within-host, mechanistic model of *P. falciparum* infection developed by Johnston *et al.*[23,24] and considered how infectiousness differs with and without antimalarial drug treatment. Because the timing of treatment relative to infection on day $t_i$ affects the net infectiousness of the primary individual, we followed previous work[25] and allowed the lag between symptom onset and treatment to vary from 0-14 days. We then compute the expected probability, $c(t_j|\ t_i)$, of transmission to a blood-feeding mosquito on day $t_j$ across each of these 15 scenarios, weighting each scenario by the probability that the lag between symptom onset and treatment was that number of days.

Because transmission of gametocytes from a primary infection depends upon mosquito blood-feeding, the expected number of mosquitoes infected on each day $t_j$ is sensitive to temporal fluctuations in mosquito density and biting rate. To account for the effects of seasonality in transmission, we multiplied each probability of transmission on day $t_j$ by the mosquito-to-human ratio, $m(t_j)$, on that day, which we modeled as

$$m(t_j) = e^{s_{secular}(t_j)s_{seasonal}(t_j)}. \quad \text{(S29)}$$

In eq. (S29), $s_{secular}(t)$ characterized the long-term trend in the mosquito-to-human ratio and was modeled as a cubic spline with two knots, and $s_{seasonal}(t)$ characterized the seasonal trend in the mosquito-to-human ratio and was modeled as a cosine function with a period of 52 weeks. We exponentiated the product of the seasonal and secular trends to ensure that the mosquito-to-human ratio was restricted to positive, real values. Because transmission from a primary infection requires blood-feeding by mosquitoes, the quantity $m(t_j)$ must be scaled by the proportion of mosquitoes, $p(t_j)$, that are blood-feeding on day $t_j$. The proportion of blood-feeding mosquitoes depends upon the biting rate, $a(T(t_j))$, and the human blood index (HBI). To compute the proportion of blood-feeding mosquitoes, we related $p(t_j)$ to $a(T(t_j))$ by

$$p(t_j) = 1 - e^{-a(T(t_j))}, \quad \text{(S30)}$$

where $T(t_j)$ is the temperature (°C) on day $t_j$ and $a(T)$ is a fitted relationship between the biting rate of *Anopheles pseudopunctipennis* and temperature.[31] Because we lacked an equivalent fitted relationship for *An. darlingi*, the dominant vector in Bolívar, we assumed that the thermal response of biting for *An. darlingi* was the same as the thermal response of biting for *An. pseudopunctipennis.* We then multiplied this quantity by the HBI (0·6875) to only account for bloodmeals that were taken on human hosts.[32] Finally, the expected number of blood-feeding mosquitoes infected on day $t_j$ by a primary infection that began on day $t_i$ is equal to

$$n_{HM}(t_j|t_i) = c(t_j|t_i)m(t_j)p(t_j)HBI. \quad (S31)$$

*Onset of Mosquito Infectiousness*

Following a bloodmeal on day $t_j$ from a primary infection, a mosquito must survive the extrinsic incubation period (EIP) to become infectious and contribute to onward transmission. We therefore calculated the proportion, $p_{EIP}(t_k|t_j)$, of mosquitoes that survived to complete the EIP on day $t_k$ following a bloodmeal on day $t_j$.

To generate this time-varying quantity, we first considered the length of the EIP. Because the EIP of malaria depends upon temperature, we followed Detinova *et al.*[33] and modeled the length of the EIP as

$$EIP\big(T(t_k)\big) = \frac{111}{T(t_k) - 16}, \quad (S32)$$

where $T(t_k)$ is the temperature (°C) on day $t_k$. To account for the effect of diurnal temperature variation on the length of the EIP, we adopted the time-varying approach taken by Siraj *et al*.[34] We calculated the temperature-dependent parasite rates, $PDR(T(t_k)) = EIP\big(T(t_k)\big)^{-1}$. This quantity represents the proportion of the EIP completed on day $t_k$. We then defined the time-varying hazard for parasite development within the mosquito as

$$h_{EIP}(t_k) = 1 - e^{-PDR(T(t_k))\Delta t}, \quad (S33)$$

In eq. (S33), $\Delta t$ is the time interval over which the hazard is defined and was set to one day. Given that a mosquito took a bloodmeal on day $t_j$, the probability that the mosquito has not completed the EIP on or before day $t_k$ is

$$S_{EIP}\big(t_k \big| t_j\big) = e^{-\sum_{t_j}^{t_k} h_{EIP}(T(t))}, \quad (S34)$$

for $t_k > t_j$. In this formulation, eq. (S34) did not account for mosquito mortality and instead assumed that all mosquitoes survived to complete the EIP. To account for the effect of mosquito mortality on the proportion of mosquitoes surviving to complete the EIP, we used a fitted relationship between mosquito mortality $\mu(T(t_k))$ and temperature (°C) for *An. gambiae*, a vector of *P. falciparum* in sub-Saharan Africa.[31] We lacked an equivalent fitted relationship for *An. darlingi*, so we assumed that the thermal response of mosquito mortality for *An. darlingi* was equivalent to the thermal response of mosquito mortality for *An. gambiae.* We then calculated the time-varying hazard of mortality as

$$h_\mu(t_k) = 1 - e^{-\mu(T(t_k))\Delta t}. \quad (S35)$$

Given that a mosquito took a bloodmeal on day $t_j$, the probability of surviving to day $t_k$ is

$$S_\mu\big(t_k \big| t_j\big) = e^{-\sum_{t_j}^{t_k} h_\mu(T(t))}, \quad (S36)$$

for $t_k > t_j$. Using eqs. (S34) and (S36), we then calculated the proportion, $p_{EIP}\big(t_k \big| t_j\big)$, of mosquitoes that survived to complete the EIP on day $t_k$ after taking a bloodmeal on day $t_j$ as

$$p_{EIP}\big(t_k \big| t_j\big) = S_\mu\big(t_k \big| t_j\big)\Big(S_{EIP}(t_k + \Delta t | t_j) - S_{EIP}\big(t_k \big| t_j\big)\Big). \quad (S37)$$

<u>*Mosquito-to-Human Transmission*</u>

After surviving the EIP, a mosquito is infectious and can transmit *P. falciparum* sporozoites to cause secondary infections. We calculated the expected number of secondary infections, $n_{MH}(t_l | t_k)$, that occurred on day $t_l$ after completing the EIP on day $t_k$.

To generate this time-varying quantity, we used eqs. (S35) and (S36) to calculate the time-varying hazard of mosquito mortality and the probability of surviving to day $t_l$ following completion of the EIP on day $t_k$. We then computed the expected number of secondary infections on day $t_l$ for each mosquito that completed the EIP on day $t_k$ as

$$n_{MH}(t_l | t_k) = S_\mu(t_l | t_k) HBI p(t_l) b, \quad (S38)$$

where HBI is the human blood index, $p(t_l)$ is the proportion of mosquitoes that are blood-feeding on day $t_l$, and $b$ is the probability of transmission from mosquitoes to humans, which we set to 0·25.[27] It is important to note that we expect that the choice of $b$ would not change the magnitude of the time-varying reproduction number that we estimate, because any misspecification of $b$ would be compensated for in the model-fitting process through our estimate of the mosquito-to-human ratio.

<u>*Reproduction Number*</u>

The above quantities represent the different components of the malaria transmission cycle. Using these quantities, we computed the expected number of secondary infections, $\boldsymbol{R}(t_l|t_i)$, on day $t_l$ caused by each primary infection that began on day $t_i$ as

$$\boldsymbol{R}(t_l|t_i) = \sum_{t_j}\sum_{t_k} n_{MH}(t_l|t_k)p_{EIP}(t_k|t_j)n_{HM}(t_j|t_i). \quad \text{(S39)}$$

Because cases are aggregated on a weekly basis, but the reproduction number was defined on a daily basis, we transformed eq. (S39) onto a weekly timescale as

$$\boldsymbol{R}(w_l|w_i) = \frac{1}{28}\sum_{t_i=7(w_i-1)}^{7w_i}\sum_{t_l=7(w_l-1)}^{7w_l}\boldsymbol{R}(t_l|t_i), \quad \text{(S40)}$$

where $w_l$ and $w_i$ denote the timing in weeks of the primary and secondary infections, respectively.

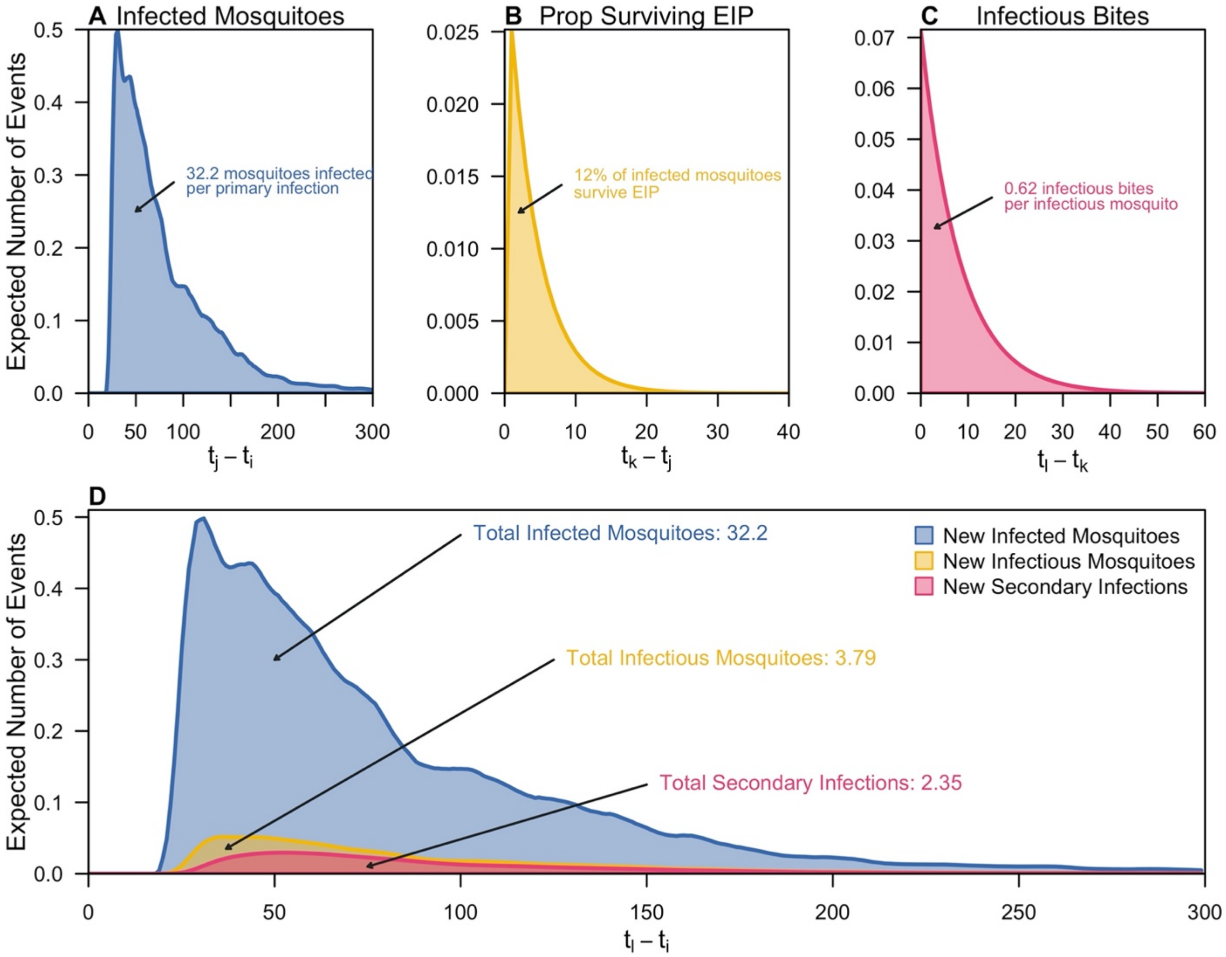


**Figure S3. Schematic of the process used to compute the time-varying reproduction number.** (A) Following the infection of a primary individual, mosquitoes are infected according to the probability of human-to-mosquito transmission, shown here for an untreated individual. (B) Upon taking an infectious bloodmeal, the probability that a mosquito survives to complete the extrinsic incubation period (EIP) is calculated. (C) After completing the EIP, the mosquito is now infectious and can transmit to a secondary mosquito. The expected number of infectious mosquito bites depends upon the probability of mosquito-to-human transmission and the mosquito mortality rate. (D) Summing the components described in (A-C) results in estimates of the number of new infected mosquitoes (blue), new infectious mosquitoes, and new secondary infections as a function of the days elapsed since the primary infection.

***Intervention Models***

*Long-Lasting Insecticidal Nets (LLINs)*

We followed previous work by Griffin *et al.*[35] and White *et al.*[27] by modeling the impact of LLINs on *P. falciparum* and *P. vivax* transmission through its effects on the length of the gonotrophic cycle, mosquito mortality, and the size of the mosquito population.

<u>*Effect on the Gonotrophic Cycle*</u>

To predict the effect of the distribution of LLINs at coverage $\chi_{LLIN}$, we first calculated its effect on the length of the gonotrophic cycle. Specifically, for a baseline human biting rate $a_0$ in the absence of LLINs ($\chi_{LLIN} = 0$), the baseline gonotrophic cycle $\delta_0$, computed as the reciprocal of human biting rate, can be subdivided into its components as

$$\delta_0 = \tau_{searching}(0) + \tau_{resting}. \quad \text{(S41)}$$

In eq. (S41), $\tau_{searching}(0)$ is the duration of time that is spent searching for a bloodmeal when $\chi_{LLIN} = 0$, and $\tau_{resting}$ is the duration of time spent resting following a bloodmeal. We assumed that LLINs prevent *An. darlingi* from taking a bloodmeal and thereby increasing the time spent searching for a bloodmeal. Accordingly, we calculated $\tau_{searching}$ as a function of LLIN coverage $\chi_{LLIN}$ as

$$\tau_{search}(\chi_{LLIN}) = \frac{\tau_{searching}(0)}{1-z}, \quad \text{(S42)}$$

where $z$ is the probability that an *An. darlingi* mosquito must begin a new search following each attempt to take a bloodmeal. This probability is calculated as

$$z = \chi_{LLIN}\phi_{LLIN}r_{LLIN}. \quad \text{(S43)}$$

In eq. (S43), $\phi_{LLIN}$ is the proportion of bites that occur while individuals are in bed, and $r_{LLIN}$ is the probability that a mosquito is repelled by a LLIN (Table S6). Taking into account the effect of LLINs on the time spent searching for a bloodmeal, the length of the gonotrophic calculated as a function of LLIN coverage is

$$\delta(\chi_{LLIN}) = \tau_{searching}(\chi_{LLIN}) + \tau_{resting}. \quad \text{(S44)}$$

The effect of LLINs on the human biting rate can therefore be computed by taking the reciprocal of eq. (S44).

<u>*Effect on the Mortality Rate*</u>

LLINs also increase the mortality of mosquitoes that come into contact with them. To quantify the effect of LLINs on mosquito mortality, we first calculated the probability that a mosquito successfully takes a bloodmeal as

$$w = \chi_{LLIN}(1 - \phi_{LLIN} + \phi_{LLIN}s_{LLLIN}) + (1 - \chi_{LLIN}), \quad \text{(S45)}$$

where $\phi_{LLIN}$ is the proportion of bites that occur while individuals are in bed, and $s_{LLIN}$ is the probability that a mosquito successfully feeds on an individual if that individual is protected by a LLIN. The probability that a mosquito survives the length of the gonotrophic cycle is equal to the probability $p_{searching}$ that the mosquito survives the searching period and the probability $p_{resting}$ that the mosquito survives the resting period. The probability that the mosquito survives the search period is affected by the coverage of LLINs and can be calculated as

$$p_{searching}(\chi_{LLIN}) = \frac{p_{searching}(0)w}{1 - zp_{searching}(0)}, \quad \text{(S46)}$$

where $p_{searching}(0)$ is the probability that a mosquito survives the searching period in the absence of LLINs, $w$ is the probability that a mosquito successfully takes a bloodmeal and is calculated in eq. (S45), and $z$ is the probability that a mosquito must begin a new search following each attempt to take a bloodmeal and is calculated in eq. (S43). Furthermore, we calculated the probability that a mosquito survives the resting period as

$$p_{resting} = e^{-\mu\tau_{resting}}. \quad \text{(S47)}$$

In eq. (S47), μ is the baseline mosquito mortality rate, and $\tau_{resting}$ is the duration of time that is spent resting following a bloodmeal. Finally, the daily mortality rate computed as a function of LLIN coverage is

$$\mu(\chi_{LLIN}) = -a(\chi_{LLIN})\log\big(p_{searching}(\chi_{LLIN})p_{resting}\big), \quad \text{(S48)}$$

where $a(\chi_{LLIN})$ is the human biting rate computed as function of LLIN coverage.

*Effect on the Mosquito Population Size*

By increasing the duration of the gonotrophic cycle and elevating the mosquito mortality rate, LLINs also affect the size of the mosquito population. To quantify this effect, we first computed the size, $M_t$, of the mosquito population in the absence of LLINs as the production of the mosquito-to-human ratio $m(t)$ and the human population size $N_t$. Following Cavany *et al.*,[36] the rate of change of the size of the mosquito population is defined as

$$\frac{dM}{dt} = \psi(t) - \mu(t)M_t, \quad \text{(S49)}$$

where $\psi(t)$ and $\mu(t)$ are the respective mosquito emergence and mortality rates at time $t$. Given estimates of $M_t$ and $\mu(t)$ at each point in time, we solved for the left-hand side of eq. (S49) numerically and rearranged the terms to calculate the mosquito emergence rate. We then computed the effect of LLINs on the size of the mosquito population as

$$M_{LLIN,t} = \psi(t) + (1 - \mu(t, \chi_{LLIN}))M_{LLIN,t-1}. \quad \text{(S50)}$$

In eq. (S50), $\mu(t, \chi_{LLIN})$ is the mosquito mortality rate at time $t$ and LLIN coverage $\chi_{LLIN}$.

*Waning in Usage and Insecticidal Activity*

Following White *et al.*,[27] we modeled the waning in LLIN usage and the activity of the insecticide during the intervention period. Given an initial target coverage $\chi_{LLIN}$, we calculated the LLIN coverage on week $t$ as

$$\chi_{LLIN}(t) = \chi_{LLIN}e^{-t/T_{LLIN}}, \quad \text{(S51)}$$

where $T_{LLIN}$ is the mean duration of LLIN usage, which we set to 84·2 weeks (Table S6). The previous equations calculated the effects of LLINs on the length of the gonotrophic cycle, mosquito mortality, and the size of the mosquito population under the assumption that the probabilities of repellency $r_{LLIN}$ and death $d_{LLIN}$ due to the insecticide do not change over time. To account for waning in insecticidal activity over time, we modeled these quantities as

$$r_{LLIN}(t) = r_{LLIN,0} + \big(r_{LLIN} - r_{LLIN,0}\big)e^{-t/\gamma_{LLIN}}, \quad \text{(S52)}$$

$$d_{LLIN}(t) = d_{LLIN}e^{-t/\gamma_{LLIN}}. \quad \text{(S53)}$$

In eqs. (S52) and (S53), $\gamma_{LLIN}$ is the mean duration of pyrethroid insecticidal activity, which we set to 130 weeks.[27] The time-varying probability that a mosquito successfully fed on an individual that is protected by a LLIN was therefore calculated as

$$s_{LLIN}(t) = 1 - r_{LLIN}(t) - d_{LLIN}(t). \quad \text{(S54)}$$

**Table S6. Parameters characterizing LLIN interventions.**

| Parameter | Description | Value | Source |
|---|---|---|---|
| $T_{LLIN}$ | Mean duration (weeks) of LLIN usage | 84·2 | 27 |
| $\gamma_{LLIN}$ | Mean duration (weeks) of insecticidal activity | 130 | 27 |
| $r_{LLIN}$ | Probability of repellency due to insecticide | 0·6 | 27 |
| $r_{LLIN,0}$ | Probability of repellency due to physical barrier | 0·2 | 27 |
| $d_{LLIN}$ | Probability of death due to insecticide | 0·3 | 27 |
| $s_{LLIN}$ | Probability of successful bloodmeal | 0·1 | 27 |
| $\phi_{LLIN}$ | Proportion of bites that occur while individuals are in bed | 0·66 | 12 |

***Mass Drug Administration (MDA)***

Following White *et al.*[27] and Brady *et al.*,[37] we modeled the impact of mass drug administration (MDA) on *P. falciparum* and *P. vivax* transmission by assuming that a target coverage $\chi_{MDA}$ of the population received treatment, irrespective of whether they were infected with *Plasmodium* spp. parasites. The target coverage, the antimalarial drugs and the level of adherence were varied to project the impact under different hypothetical MDA programs. The modeled effects of different antimalarial drugs are considered below.

*Artemisinin Combination Therapy (ACT)*

For MDA targeting *P. falciparum* transmission, we assumed that individuals received artemisinin combination therapy (ACT). The action of the ACT depended upon whether or not the individual was infected at the point of enrollment in the MDA. For uninfected individuals, we assumed, by default, that treatment with ACT provided three weeks of prophylaxis.[35] For individuals infected with *P. falciparum* and not already receiving treatment at the time of MDA, we assumed that ACT cleared their infection, thereby reducing the reservoir of *P. falciparum* infections.

Following WHO guidelines,[38] treatment with ACT is contraindicated in pregnant women during the first trimester. Accordingly, we assumed that the MDA would screen for pregnant women and prevent them from receiving treatment. We calculated the annual number of pregnancies in Bolívar according to population-weighted estimates from WorldPop[39] and multiplied that quantity by 0·25 years, the duration of the first trimester, to arrive at the number of first-trimester pregnancies $PR_{1TM}$ at any given point in the year. The effective coverage of the MDA was then calculated as

$$\chi'_{MDA} = \left(1 - \frac{PR_{1TM}}{N_t}\right)\chi_{MDA}. \quad \text{(S55)}$$

Eq. (S55) can be interpreted as the production of the target coverage by the fraction of individuals in the population who are not in their first trimester of pregnancy.

*Chloroquine and Primaquine (CQ+PQ)*

For MDA targeting *P. vivax*, we assumed that chloroquine (CQ) and primaquine (PQ) would be used. In infected individuals, CQ clears the bloodstream *P. vivax* infection, and a 14-day (or 7-day double dose) course of PQ targets the liver-stage hypnozoites. In uninfected individuals, we assumed that treatment with CQ+PQ provided prophylaxis for 14 days.[27] As with MDA with ACT, we assumed that individuals already receiving treatment for *P. vivax* infection would not be enrolled.

Radical cure with PQ is challenging, because it can cause acute hemolysis in individuals with glucose-6-phosphate (G6PD) deficiency.[38,40] Per WHO guidelines,[38] we assumed that no G6PD screening occurred as part of the MDA but that monitoring of primaquine-induced hemolysis was performed. Under this assumption, G6PD-deficient individuals would be forced to discontinue PQ treatment and would not clear all hypnozoites from the liver. We calculated the prevalence, $f_{G6PD}$, of G6PD-deficient individuals in Bolívar according to population-weighted estimates from the Malaria Atlas Project.[7,41]

As with ACT, PQ is contraindicated in pregnant women.[38] Accordingly, we computed the effective coverage of MDA with CQ+PQ as

$$\chi'_{MDA} = \left(1 - \frac{PR_t}{N_t}\right)\chi_{MDA}, \quad \text{(S56)}$$

where $PR_t$ is the number of pregnancies at time *t*.

#### Inference Framework

We used a Bayesian inference framework to fit our *P. falciparum* and *P. vivax* transmission models to weekly incidence and annual prevalence data. This framework allowed us to quantify uncertainty in our estimates and propagate this uncertainty into our projections of intervention impact.

##### *Likelihoods*

The core of our inference framework is the likelihood, which is the probability of the data given the model and a particular set of parameters. Because we integrated multiple data types into our inference framework, we calculated the overall likelihood as the product of the likelihood associated with each data type. The data types used were weekly *P. falciparum* autochthonous and recrudescing cases, weekly *P. vivax* autochthonous and relapsing cases, and annual *P. vivax* prevalence.

<u>Incidence Data</u>

The transmission models track the weekly number of reported autochthonous and recrudescing *P. falciparum* cases and the weekly number of reported autochthonous and relapsing *P. vivax* cases. We specified the likelihood of the incidence data reported by the Venezuelan Ministry of Health in Bolívar given the incidence predicted by the models as

$$\mathcal{L}\left(\vec{I}_{Pf}, \vec{\xi}_{Pf}, \vec{I}_{Pv}, \vec{\xi}_{Pv} \middle| \vec{T}^{(I)}, \vec{T}^{(\xi,c)}, \vec{T}^{(\xi,m)}, \vec{T}_0^{(c)}, \vec{T}_0^{(m)}, \vec{T}_L^{(c)}, \vec{T}_L^{(m)}, \vec{T}_\xi^{(c)}, \vec{T}_\xi^{(m)}\right)$$
$$= \prod_t \text{Negative Binomial}\left(I_{Pf,t} \middle| T_t^{(I)} + T_t^{(\xi,m)}, \kappa_{I_{Pf}}\right) \text{ x Negative Binomial}\left(\xi_{Pf,t} \middle| T_t^{(\xi,c)}, \kappa_{\xi_{Pf}}\right) \text{ x}$$
$$\text{Negative Binomial}\left(I_{Pv,t} \middle| T_{0,t}^{(c)} + T_{L,t}^{(c)} + T_{\xi,t}^{(m)}, \kappa_{I_{Pv}}\right) \text{ x Negative Binomial}\left(\xi_{Pv,t} \middle| T_{0,t}^{(m)} + T_{L,t}^{(m)} + T_{\xi,t}^{(c)}, \kappa_{\xi_{Pv}}\right). \quad (S57)$$

In eq. (S57), $\vec{I}_{Pf}$ and $\vec{\xi}_{Pf}$ are the respective weekly reported number of autochthonous and recrudescing *P. falciparum* cases, and $\vec{I}_{Pv}$ and $\vec{\xi}_{Pv}$ are the respective weekly reported number of autochthonous and relapsing *P. vivax* cases. The model predictions for each of these quantities are provided in eq. (S57), and a full description of the state variables can be found in Tables S2 & S4. For each data type, the dispersion parameter for the negative binomial distribution was chosen by through maximum-likelihood estimation of that data type given the mean of the negative binomial distribution at each time point as the cubic smoothing spline of the data with four knots. The dispersion parameter was 5.76 for *P. falciparum* autochthonous cases, 3.47 for *P. falciparum* recrudescing cases, 9.02 for *P. vivax* autochthonous cases, and 7.85 for *P. vivax* relapsing cases. Given the high number of parameter estimated and the challenges associated with obtaining convergence, we chose to estimate this parameter independent of model fitting.

A complete record of the weekly number of reported autochthonous and recrudescing *P. falciparum* cases and the weekly number of reported autochthonous and relapsing *P. vivax* cases was available during 2000-2017, and a partial record was available for 2018. The annual number of autochthonous malaria cases aggregated across *Plasmodium* spp. was available for 2018 and 2019. We therefore specified a negative binomial likelihood for the annual number of autochthonous malaria cases reported by the Venezuelan Ministry of Health in Bolívar given the annual number of autochthonous malaria cases predicted by the model.

<u>Prevalence Data</u>

The transmission models also predicted the weekly prevalence of *P. falciparum* and *P. vivax*. However, age-independent estimates of prevalence were only available on an annual basis for *P. vivax*.[6] To relate these quantities, we computed the mean annual prevalence predicted by our transmission models as the average of the weekly predicted prevalence estimates. We then specified the marginal likelihood of the prevalence data $\vec{p}_{Pv}$ given the mean annual prevalence $\vec{\bar{p}}_{Pv}$ predicted by the model as

$$\mathcal{L}\left(\vec{p}_{Pv} \middle| \vec{\bar{p}}_{Pv}, \vec{\bar{N}}\right) = \prod_{2000}^{2017} \left( \frac{1}{11} \sum_{m=1}^{11} \text{Binomial}\left( \lfloor \bar{N}_{m,t} p_{Pv,m,t} + 0 \cdot 5 \rfloor \middle| \bar{N}_{m,t}, \bar{p}_{Pv,t} \right) \right), \quad (S58)$$

where $\bar{N}_{m,t}$ is the mean population size in municipality *m* during year *t* rounded to the nearest integer.

##### *Parameters*

By fitting our model to weekly case counts and annual prevalence estimates, we estimated the parameters of the *P. falciparum* and *P. vivax* transmission models. A full description of the parameters that were estimated and their prior ranges can be found in Table S7. For the self-medication rate, a beta-prior for each year was constructed, based upon estimates from personnel in Bolívar. Unless specified otherwise, the prior distribution placed on each parameter was uniform within the prior range.

**Table S7. Parameters estimated during model-fitting**

| Parameter | Description | Species | Prior | Source |
|---|---|---|---|---|
| $k_m^{(1)}, k_m^{(2)}$ | Knots for secular component of mosquito-to-human ratio | Same for *Pf* and *Pv* | [0,10] | |
| $A$ | Amplitude of seasonal component of mosquito-to-human ratio | Same for *Pf* and *Pv* | [0,2·5] | |
| $t_{peak}$ | Peak week of seasonal component of mosquito-to-human ratio | Same for *Pf* and *Pv* | [0,51] | |
| $\theta_{PQ}$ | Proportion receiving PQ treatment | *Pv* only | [0,1] | |
| $\theta_{ACT}$ | Proportion adhering to ACT | *Pf* only | [0,1] | |
| $\sigma_R$ | Reproduction number multiplier for *P. vivax* | *Pv only* | [0,10] | |
| $I_0$ | Initial weekly number of infections | Separate for *Pf* and *Pv* | [1,10000] | |
| $k_\rho^{(1)}, k_\rho^{(2)}$ | Knots for *P. falciparum* reporting rate | *Pf* only | [-3,0] | |
| $k_\rho^{(1)}, k_\rho^{(2)}, k_\rho^{(3)}$ | Knots for *P. vivax* reporting rate | *Pv* only | [-3,0] | |
| $\chi(t)$ | Self-medicating rate | Separate for *Pf* and *Pv* | [0,1] | |
| $\phi_{D,Min}$ | Minimum probability of developing *P. vivax* symptoms | *Pv only* | [0,0·22] | [27] |
| $\phi_{D,Max}$ | Maximum probability of developing *P. vivax* symptoms | *Pv only* | [0·61, 0·983] | [27] |

***Sequential Monte Carlo (SMC)***

We fitted the transmission models jointly using a sequential Monte Carlo algorithm [42]. We initialized a population of 200,000 particles by sampling values for each parameter from the respective prior distribution. For each particle, we calculated the likelihood of the model given the observed data. Particles were then weighted according to their likelihood values and a temperature parameter. The latter quantity flattened the likelihood in early iterations of the algorithm to maintain particle diversity, and its initial value was chosen such that that 90% of the particle weights were allocated to the top 20% of particles with the highest likelihood values. We adopted a cooling schedule such that the temperature parameter was 80% of its previous value at subsequent iterations. After assigning weights to each particle in the population, we resampled 200,000 particles proportional to those weights and fitted a multivariate normal distribution to the resampled particles. Finally, 200,000 particles were randomly sampled from the fitted multivariate normal distribution. This algorithm was repeated until the temperature parameter approached one, signifying that the likelihood was no longer artificially flattened. We assessed convergence by running five chains in parallel and computing the Gelman-Rubin statistic for each parameter. Gelman-Rubin statistics less than 1·1 were interpreted as providing support for convergence.[43]

**Analyses**

***Simulation Study***

The assess the extent to which our Bayesian inference framework was able to correctly estimate the parameters listed in Table S7, we performed a simulation study. We generated 10 synthetic data sets by sampling from the prior ranges specified in Table S7 and simulating under our *P. falciparum* and *P. vivax* transmission models. We then fitted the transmission models to the synthetic data sets using our SMC approach. We compared the inferred 95% credible intervals with the true values for each parameter and performed posterior predictive checks to ensure that our model captured the variation in the simulated data with sufficient uncertainty.

***Application to Epidemiological Data***

To characterize the historical patterns of transmission in Bolívar, we fitted our *P. falciparum* and *P. vivax* transmission models to the weekly case counts and cumulative incidence from the Venezuelan Ministry of Health and the annual prevalence estimates from the Malaria Atlas Project. As an independent validation of our estimates, we compared the landing catch and parity estimates obtained from the entomological component of our model to data on landing catches and parity collected in Bolívar.[13,44]

***Projecting the Impact of Interventions***

After fitting the *P. falciparum* and *P. vivax* transmission models to the epidemiological data, we considered the impact that interventions might have in the region should appropriate resources to support surveillance and control be brought to bear. For the purpose of this impact analysis, we assumed that the implementation of interventions would occur at the start of 2021. For MDA, we varied the target coverage, the antimalarial drugs, and the level of assumed adherence. For LLINs, we varied the target coverage. The target coverage for LLINs ranged from 10% to

90% in increments of 10%, and the target coverage for MDA ranged from 50% to 80% in increments of 5%. For each intervention scenario, we calculated the median posterior number of *P. falciparum* and *P. vivax* infections averted and reported the 95% posterior prediction intervals as well.

**Sensitivity Analyses**

***Impact of Spatial Heterogeneity***

Previous work has demonstrated that spatial aggregation of epidemiological data can obscure important heterogeneities that may be relevant for the transmission and control of infectious diseases.[45,46] In our primary analysis, we fitted our transmission models to state-level data. To determine whether heterogeneity at the municipality level might affect our conclusions about historical patterns of transmission and the projected impacts of interventions, we re-fitted our transmission models to data from the Sifontes municipality, the epicenter of the epidemic in Bolívar, and projected the impact of control interventions, should they be implemented in Sifontes.

***Uncertainty in LLIN Action***

In the primary analysis, we assumed values from the literature to model the effect of LLINs against mosquitoes (Table S6). However, these parameters were informed by experimental hut studies in Tanzania where the dominant vectors are *An. gambiae* and *An. funestus*.[27,47] Furthermore, estimates of LLIN usage were obtained from a study conducted in Papua New Guinea.[48] These parameters informing our projections of the impact of LLINs may not reflect the impact of LLINs on *An. darlingi*. To address this uncertainty, we performed a sensitivity analysis in which we assumed that each parameter was 50% lower than its original value that we assumed in the primary analysis (Table S6). We then re-projected the impact of interventions to quantify the impact on LLIN action on the projected impact.

***Sensitivity of Impact Projections to Inference Replicates***

In the event that our SMC chains did not convergence on the basis of the Gelman-Rubin statistic, we performed a sensitivity analysis of our impact projections. Specifically, we compared the projected impact of the intervention scenarios across the SMC chains. In doing so, we can determine whether a lack of convergence of the parameters on the basis of the Gelman-Rubin statistic affects the intervention impact conclusions that we reached.

#### Results

**Daily Temperature Estimates in Bolívar**

We estimated the daily, population-weighted temperatures in Bolívar during 1995-2021 (Fig. S4). Comparison of the predicted daily temperatures to the data collected from 157 weather stations in Venezuela, Colombia, Ecuador, Peru, Guyana, and Suriname showed strong agreement with a Pearson correlation coefficient of 0.93. The strong correlation provided evidence that we captured the temporal trends in the region, supporting use of the daily temperature estimates as inputs to our *P. falciparum* and *P. vivax* transmission models.

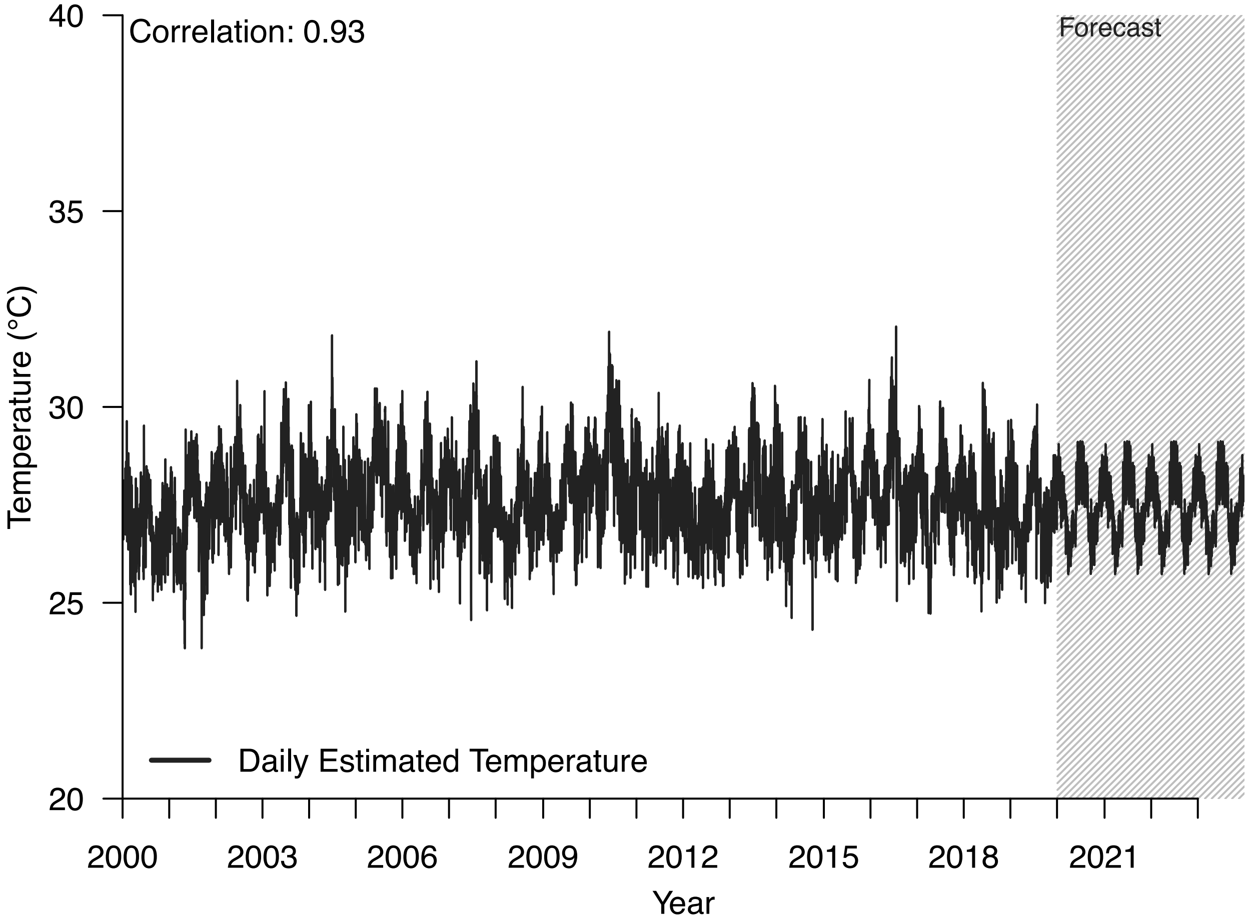


**Figure S4. Daily temperature estimates in Bolívar, Venezuela.** The time series of daily, population-weighted mean temperatures for Bolívar that were estimated using a spatial kriging approach are shown. Forecasted daily mean temperatures (shown in the shaded region) were computed as the three-year average of mean daily temperatures for each day.

**Simulation Study**

To confirm that our Bayesian inference framework could accurately estimate the parameters of the *P. falciparum* and *P. vivax* transmission models, we simulated 10 data sets using our simulation framework. We then measured how well our Bayesian inference framework recovered the true parameter values and how well we reproduced the epidemiological dynamics.

The estimated marginal posterior distributions were generally in good agreement with the true parameter values (Fig. S5). Parameters related to the proportion of individuals self-medicating were more challenging to estimate, which may reflect an inherent limitation of the data for inferring these quantities. Using the inferred parameter estimates, the posterior predictive distributions reproduced the simulated data (Fig. S5) as well as the underlying epidemiological quantities. These results indicated that the models can accurately estimate the transmission dynamics from routinely collected surveillance data, a feature needed to make reliable projections of the impact of interventions.

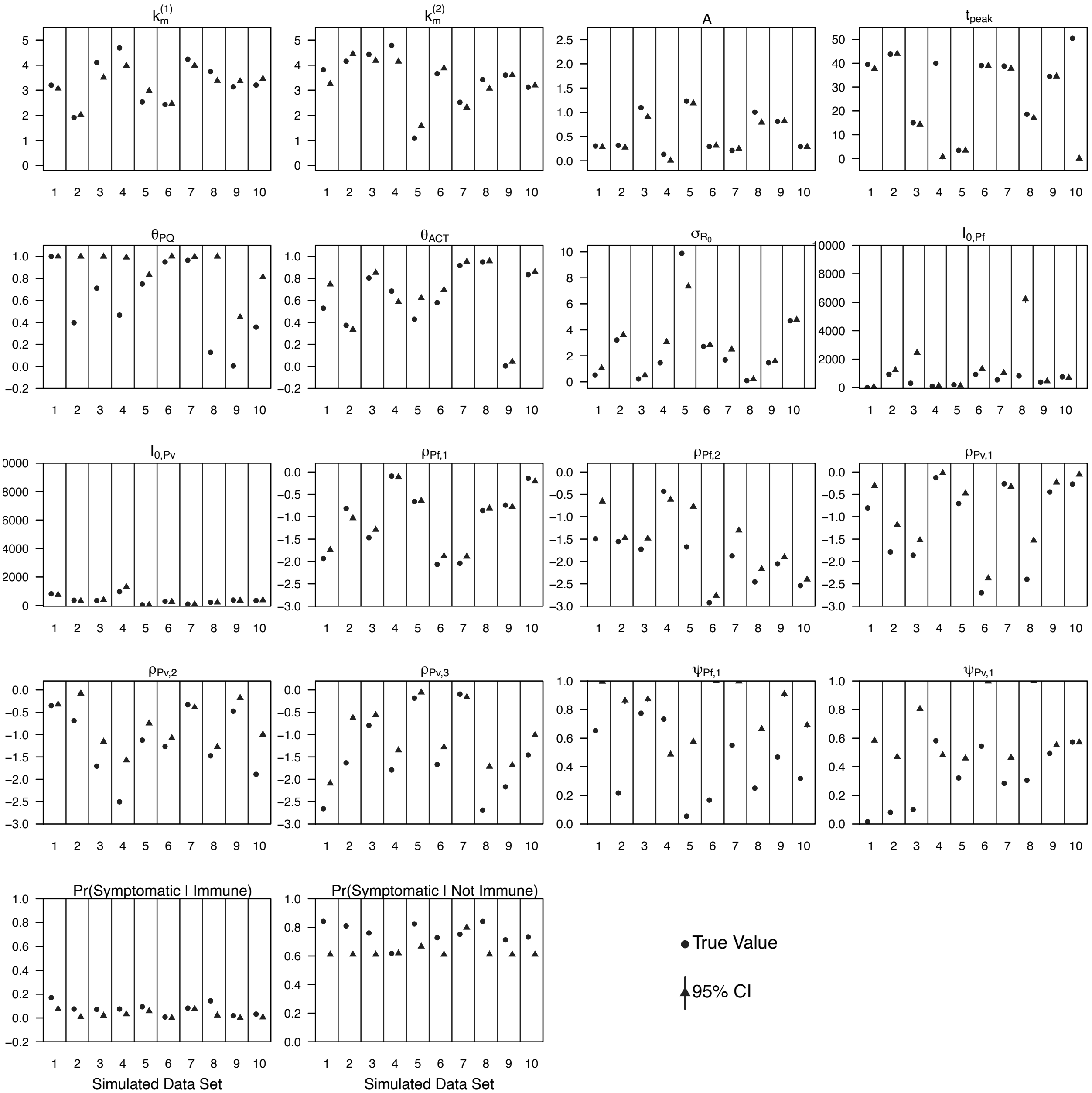


**Figure S5. Comparison of the true and estimated parameters from the simulated data sets.** The true values (circles) are compared to the posterior median (triangles) and 95% credible intervals (segments) inferred from the 10 simulated data sets for each model parameter.

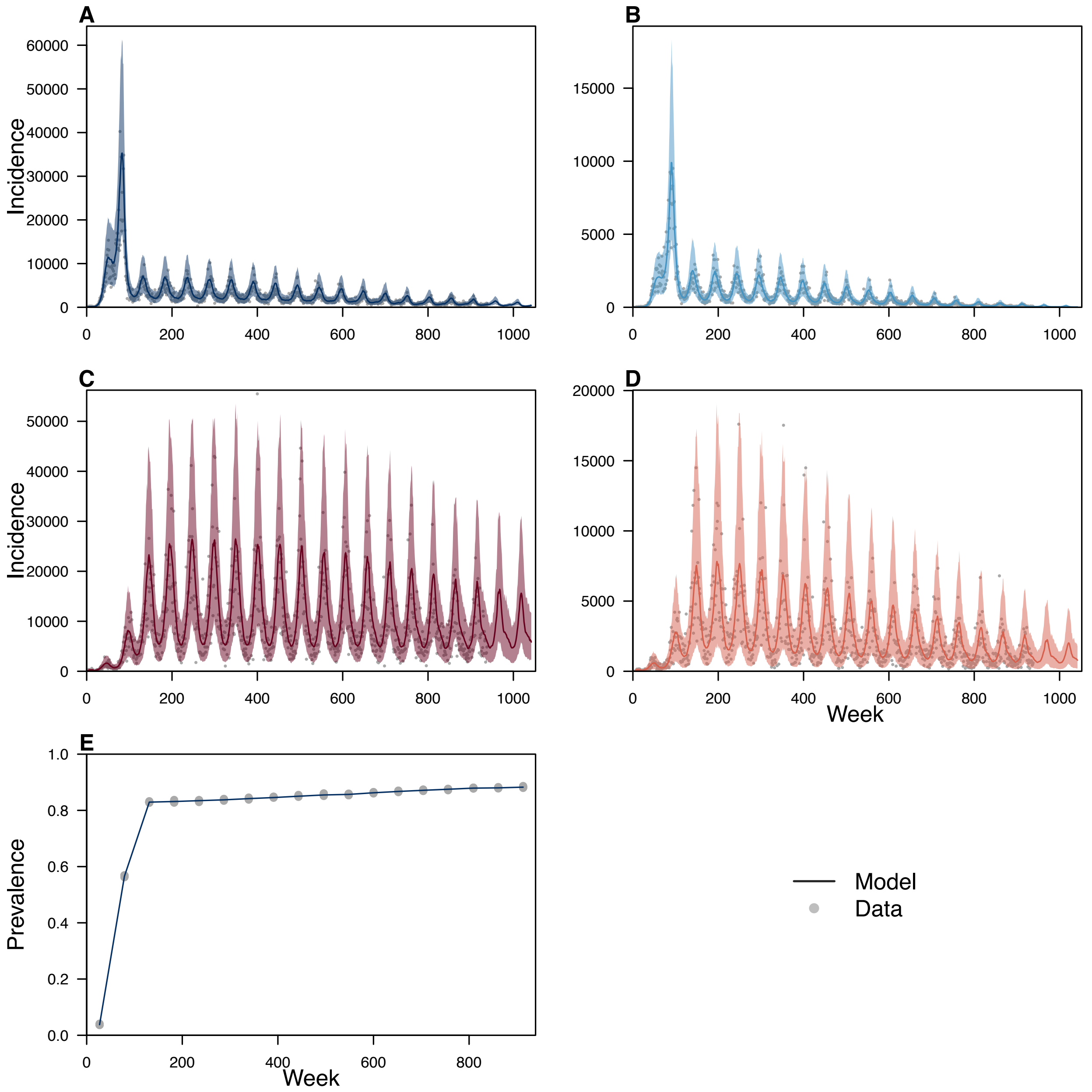


**Figure S6. Comparison of the fitted model to the simulated data.** The 95% credible interval and 95% posterior predictive intervals of the fitted transmission models are compared to the simulated incidence of (A) *P. vivax* autochthonous cases, (B) *P. falciparum* autochthonous cases, (C) *P. vivax* relapse cases, (D) *P. falciparum* recrudescence cases, and (E) *P. vivax* annual prevalence estimates. The results correspond to simulated data set #9 in Figure S5.

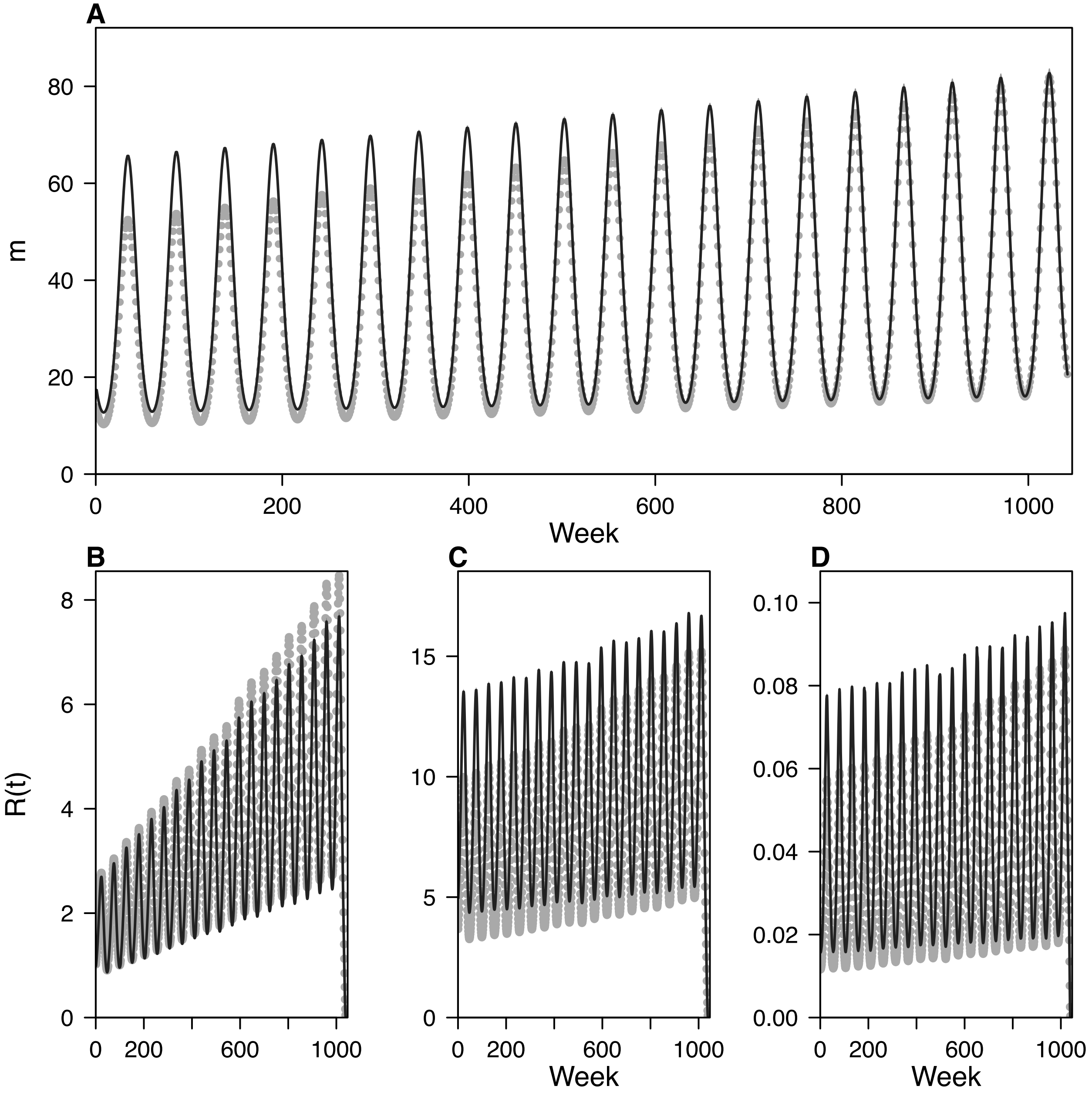


**Figure S7. Comparison of fitted and simulated mosquito-to-human ratio and reproduction numbers.** The 95% credible intervals are compared to the simulated (A) mosquito-to-human ratio, the time-varying reproduction number of (B) *P. falciparum*, and the time-varying reproduction number of *P. vivax* for (C) untreated and (D) treated infections. Dots are the true simulated quantities, and lines are the inferred posterior quantities. These results correspond to simulated data set #9 in Figure S5.

**Application to Epidemiological Data**

Following the validation exercises, we jointly fitted the *P. falciparum* and *P. vivax* transmission models to the epidemiological data collected during 2000-2019. We performed 6 independent replicates of the SMC algorithm, each with 100,000 particles per generation. Qualitatively, each independent replicate obtained similar estimates for each parameter (Fig. S8), and the ratio of the variance between the median parameter estimates relative to the variance of the prior distribution was 0.003 on average and ranged from $4.0 \times 10^{-7}$ to 0.02. This suggested that the independent replicates identified a narrow region of the multi-dimensional parameter space. Nevertheless, the Gelman-Rubin statistics exceeded 1.1 due to the narrow credible intervals for each independent replicate, indicating a lack of convergence. Therefore, we chose the independent replicate with the highest median likelihood for our primary analysis and additionally evaluated the sensitivity of our impact projections to this choice.

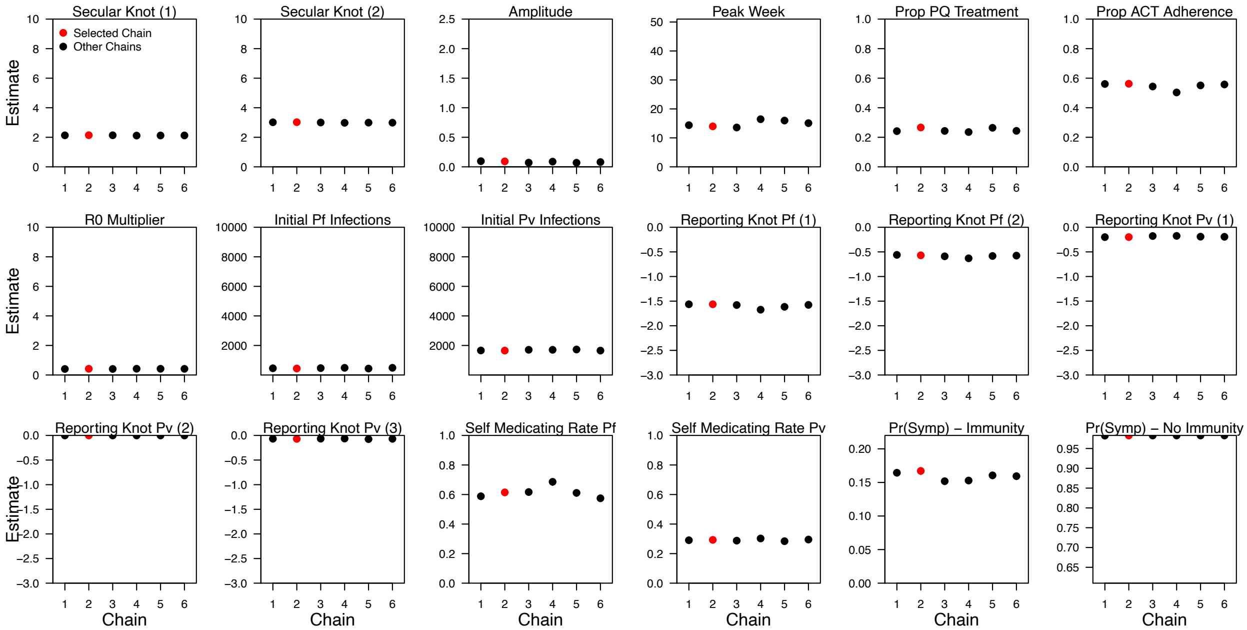


**Figure S8. Posterior parameter estimate.** The median posterior parameter estimates (points) and 95% credible intervals (bars) are compared for the selected inference replicate (red) and all other inference replicates (black).

The fitted transmission models captured the temporal patterns in incidence and prevalence of *P. falciparum* and *P. vivax* (Figs. 1, S9). We noted strong agreement between the posterior predictive distribution and the data, and the models reproduced the interannual increase in incidence as well as seasonal variation therein. In 2018 and 2019, a complete record of weekly incidence disaggregated by *Plasmodium* spp. was not available. Therefore, we fitted to cumulative malaria incidence for 2018 and 2019, and the model predictions were generally consistent with the reported cumulative incidence (Fig. S9A). We estimated that in 2018 there were 194,000 (95% PPI: 179,000 – 210,000) malaria cases, compared to the reported value of 223,404. Similarly, we estimated that in 2019 there were 244,000 (224,000 – 266,000) malaria cases, compared to the reported value of 230,035. Our fitted *P. vivax* prevalence estimates were consistent with the range of the population-weighted *P. vivax* prevalence estimates reported by Battle *et al*. during 2000-2017. It is important to note that there is uncertainty associated with the modeled geospatial estimates that we were unable to account for during model fitting given that appropriate descriptions of uncertainty are not available for those estimates. Furthermore, a recent study by Kamau *et al.*[49] compared Malaria Atlas Project estimates to surveys of clinical incidence in Kenya and identified a weak association, which serves as a reminder of the limitations of modeled quantities such as these prevalence estimates.

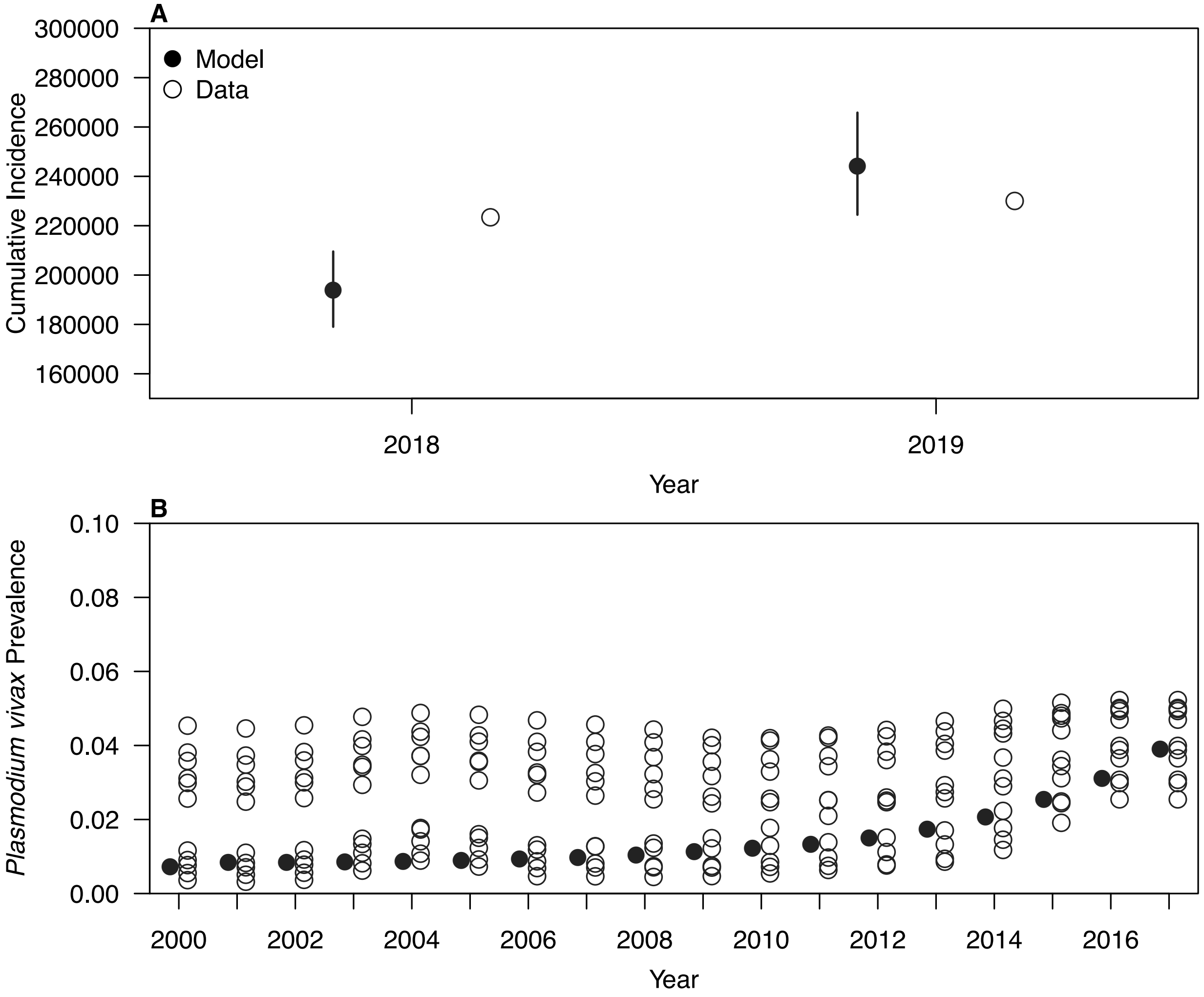


**Figure S9. Comparison of fitted models to epidemiological data.** The 95% credible intervals and the 95% posterior predictive intervals obtained from the fitted models are compared to the data for (A) annual cumulative *Plasmodium* spp. autochthonous cases and (B) annual *P. vivax* prevalence estimates from the Malaria Atlas Project.[6] In (B), each point is a prevalence estimate from a municipality in Bolívar.

The fitted model provides insight into the epidemiological dynamic of *P. falciparum* and *P. vivax* in Bolívar during 2000-2019. We estimated that a median 56.3% (55.3 – 57.2%) of individuals seeking treatment for *P. falciparum* infections in a health clinic adhered to ACT and that 26.6% (25.7 -27.6%) of individuals seeking treatment for *P. vivax* infection in a health clinic received radical cure with PQ. That the reporting rate for *P. falciparum* increased over the study period suggested that proportion of individuals seeking treatment in a health clinic for *P. falciparum* infections increased over time. By contrast, the reporting rate for *P. vivax* initially decreased and then increased, indicating that the proportion of individuals seeking treatment in a health clinic for *P. vivax* infections initially decreased and then eventually increased over time. Furthermore, we estimated that the ratio of mosquitoes to humans increased over the study period. This increase could be attributed to increases in the mosquito population within Bolívar due to an expansion of larval habitats, for instance, and may also reflect increased contact between mosquitoes and humans due to occupational hazards in the region, such as illegal gold mining.[9]

We independently validated the entomological predictions from our fitted model against data collected in the region (Fig. S10). Our model predictions for the proportion of vectors that are parous (Fig. S10B), the biting rate (Fig. S10C), and human landing catch collections (Fig. S10D) fall within the range of values reported by Moreno *et al.*[44] and Rubio-Palis *et al.*[13] We do not capture the full range of variability exhibited by the data. However, only the mosquito-to-human ratio was estimated with the model. The biting rate and the mortality rate were assumed to vary with temperature, according to a model by Mordecai *et al.*[31]

**Table S8. Joint parameter estimates for *P. falciparum* and *P. vivax* transmission models**

| Parameter | Description | Posterior Estimate |
|---|---|---|
| $k_m^{(1)}$ | 1st knot for secular component of mosquito-to-human ratio | 2.141 (2.139 – 2.144) |
| $k_m^{(2)}$ | 2nd knot for secular component of mosquito-to-human ratio | 3.022 (3.016 – 3.027) |
| $A$ | Amplitude of seasonal component of mosquito-to-human ratio | 0.0918 (0.0823 – 0.101) |
| $t_{peak}$ | Peak week of seasonal component of mosquito-to-human ratio | 13.97 (13.27 – 14.67) |

**Table S9. Parameter estimates for *P. falciparum* transmission model**

| Parameter | Description | Posterior Estimate |
|---|---|---|
| $\theta_{ACT}$ | Proportion adhering to ACT | 0.563 (0.553 – 0.572) |
| $I_0$ | Initial weekly number of infections | 439.0 (428.5 – 449.5) |
| $k_\rho^{(1)}$ | 1st knot for reporting rate | -1.564 (-1.577 – -1.551) |
| $k_\rho^{(2)}$ | 2nd knot for reporting rate | -0.569 (-0.580 - -0.559) |
| $k_\varrho^{(1)}$ | 1st knot for self-medicating rate | 0.615 (0.594 – 0.635) |

**Table S10. Parameter estimates for *P. vivax* transmission model**

| Parameter | Description | Posterior Estimate |
|---|---|---|
| $\theta_{PQ}$ | Proportion receiving PQ treatment | 0.266 (0.257 – 0.276) |
| $\sigma_R$ | Reproduction number multiplier for *P. vivax* | 0.419 (0.416 – 0.422) |
| $I_0$ | Initial weekly number of infections | 1652 (1630 – 1673) |
| $k_\rho^{(1)}$ | 1st knot for reporting rate | -0.200 (-0.205 – -0.194) |
| $k_\rho^{(2)}$ | 2nd knot for reporting rate | -2.34 (-4.76 – -0.0630) x $10^{-4}$ |
| $k_\rho^{(3)}$ | 3rd knot for reporting rate | -0.0701 (-0.0725– -0.0676) |
| $k_\varrho^{(1)}$ | 1st knot for self-medicating rate | 0.293 (0.288 – 0.297) |
| $\phi_{D,Min}$ | Minimum probability of developing *P. vivax* symptoms | 0.167 (0.162 – 0.172) |
| $\phi_{D,Max}$ | Maximum probability of developing *P. vivax* symptoms | 0.983 (0.983 – 0.983) |

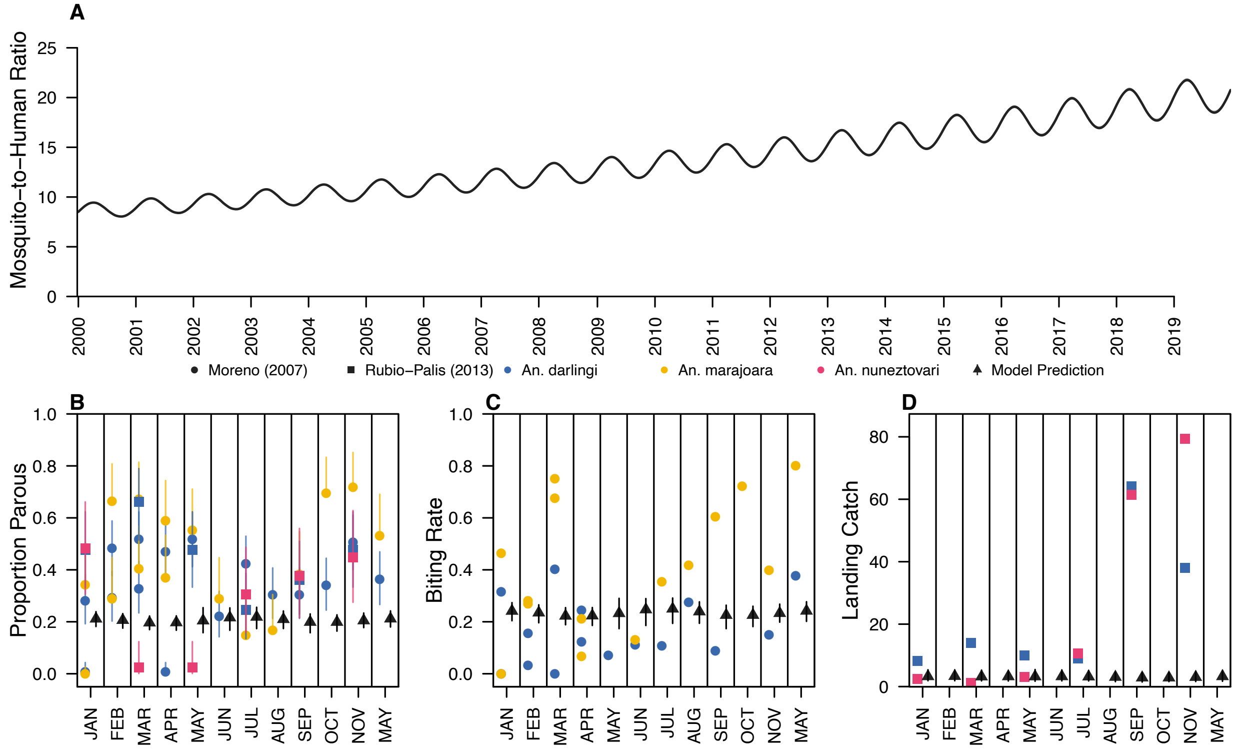


**Figure S10. Independent validation of entomological estimates.** (A) The estimated mosquito-to-human ratio over time. The model predictions (black triangles) for (B) proportion parous, (C) biting rate, and (D) human landing catch collections are compared to data collected by Moreno *et al.* (2007) (circles) and Rubio-Palis *et al.* (2013) (squares) for *An. darlingi* (blue), *An. marajoara* (yellow), and *An. nuneztovari* (pink).

#### Impact of Spatial Heterogeneity

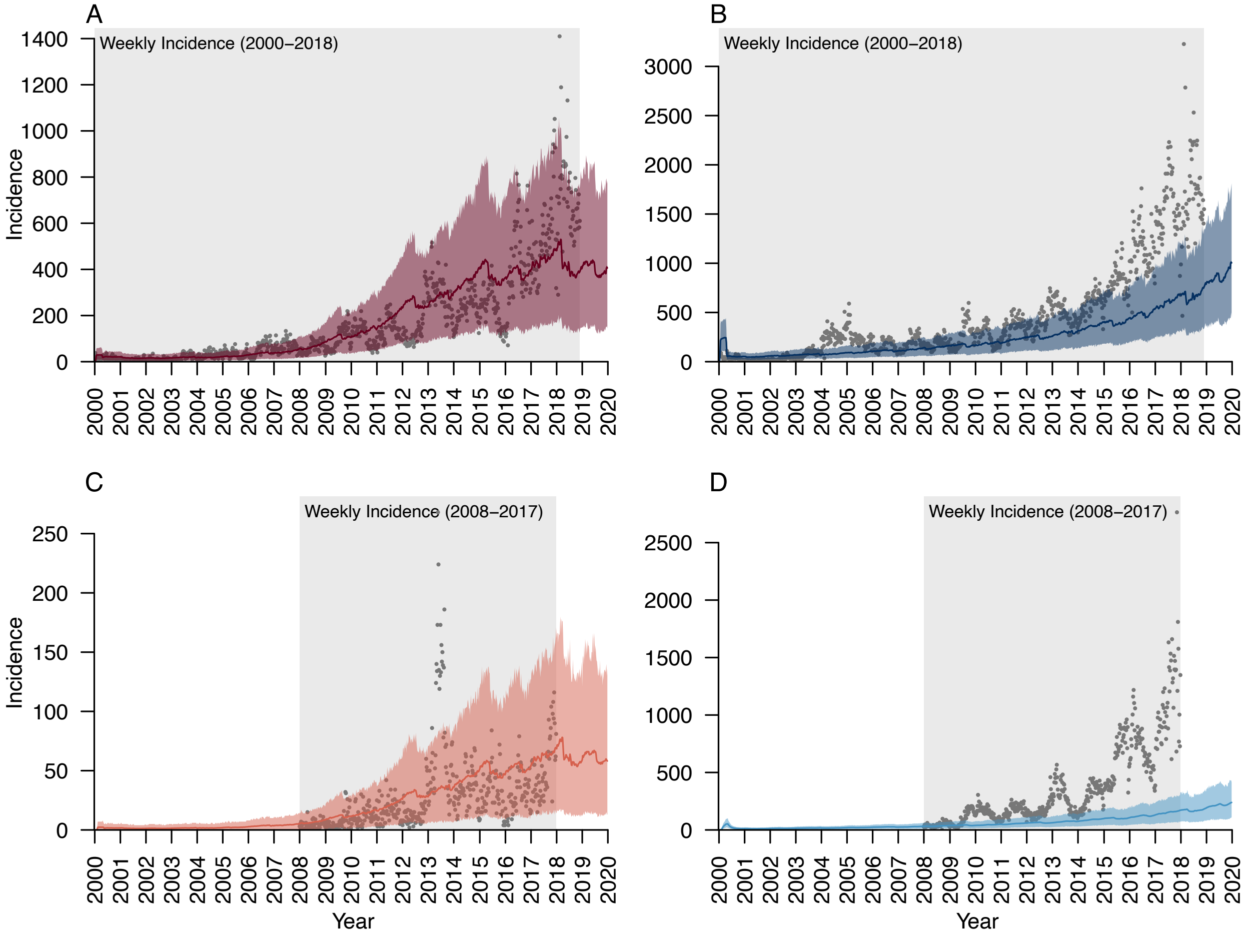


**Figure S11. Comparison of fitted transmission models to weekly epidemiological data in Sifontes.** The fitted *P. falciparum* and *P. vivax* transmission models are compared to the weekly incidence data for (A) *P. falciparum* autochthonous cases, (B) *P. vivax* autochthonous cases, (C) *P. falciparum* recrudescence cases, and (D) *P. vivax* relapse cases. The shaped area represents the period for which weekly incidence data was available, and each gray point is the reported incidence in a given week. The colored line is the median posterior estimate from the fitted transmission model, and the colored shaded region is the 95% posterior prediction interval.

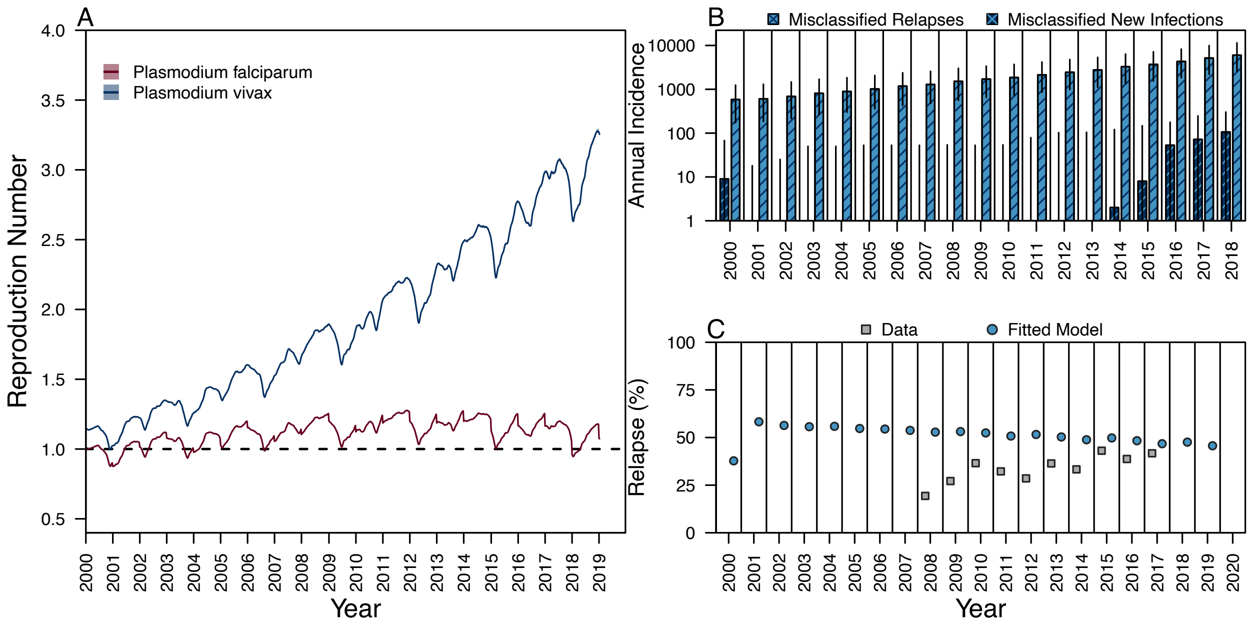


**Figure S12. Transmission and epidemiological dynamics in Sifontes.** (A) The posterior median (line) and 95% credible interval (shaded region) of the time-varying reproduction number are shown for *P. falciparum* (red) and *P. vivax (*blue). (B) The annual incidence of *P. vivax* relapses that are misclassified as new infections (light blue) and *P. vivax* new infections that are misclassified as relapses (dark blue) are shown during 2000-2018. Segments denote the 95% posterior prediction intervals. (C) The annual percentage of all *P. vivax* infections during 2000-2020 that are relapses. Each gray point is the percentage obtained from the epidemiological data (available during 2008-2017), and each blue point is the percentage of all *P. vivax* infections, both reported and unreported, that are relapses from the fitted transmission model.

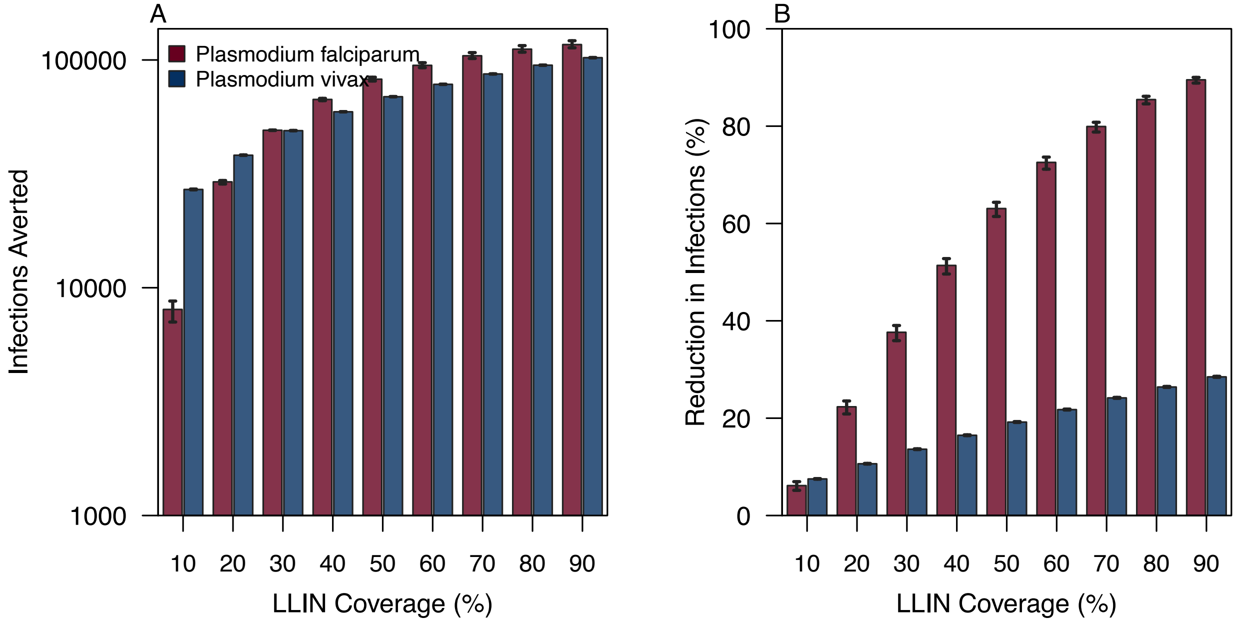


**Figure S13. Projected impact of LLIN distribution during 2021-2023 in Sifontes.** (A) The expected number of *P. falciparum* (red) and *P. vivax* (blue) infections averted during 2021-2023 is projected as a function of LLIN coverage. (B) The effect size of LLIN distribution is projected as a function of LLIN coverage. In both plots, the height of the bar denotes the median of the posterior prediction distribution, and the segments are the 95% posterior prediction interval.

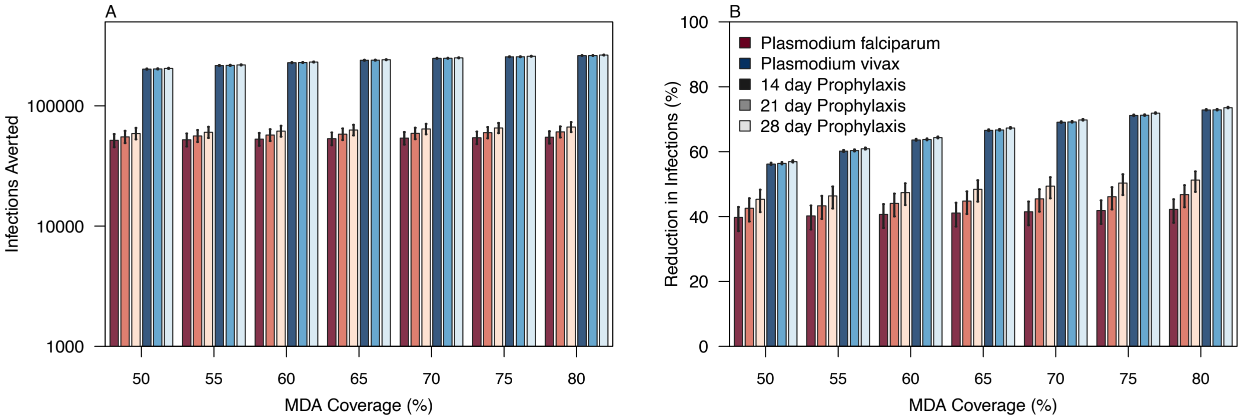


**Figure S14. Projected impact of MDA (CQ vs. ACT) during 2021-2023 in Sifontes.** (A) The expected number of *P. falciparum* (red) and *P. vivax* (blue) infections averted during 2021-2023 is projected as a function of MDA coverage for varying durations of prophylaxis. (B) The effect size of each MDA is projected as a function of coverage. Adherence to treatment is assumed to be 100%. In both plots, the height of the bar is the median of the posterior prediction distribution, and the segments are the 95% posterior prediction interval.

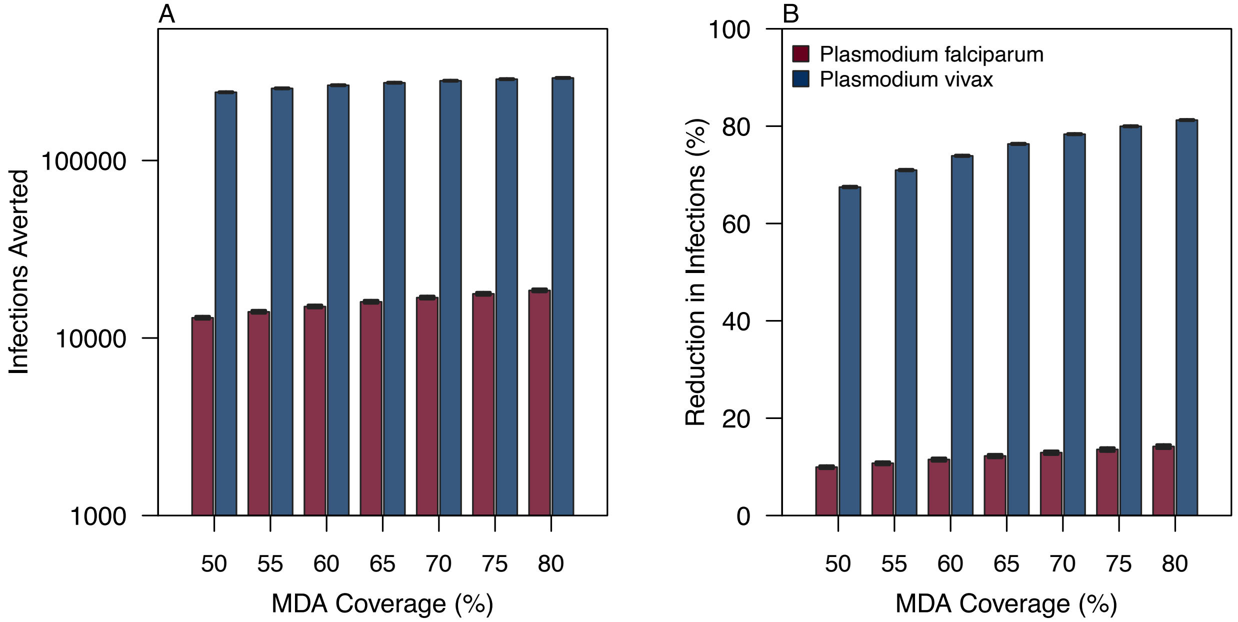


**Figure S15. Projected impact of MDA (CQ + PQ) during 2021-2023 in Sifontes.** (A) The expected number of *P. falciparum* (red) and *P. vivax* (blue) infections averted during 2021-2023 is projected as a function of MDA coverage. (B) The effect size of MDA is projected as a function of coverage. Adherence to treatment is assumed to be 100%. In both plots, the height of the bar denotes the median of the posterior prediction distribution, and the segments are the 95% posterior prediction interval.

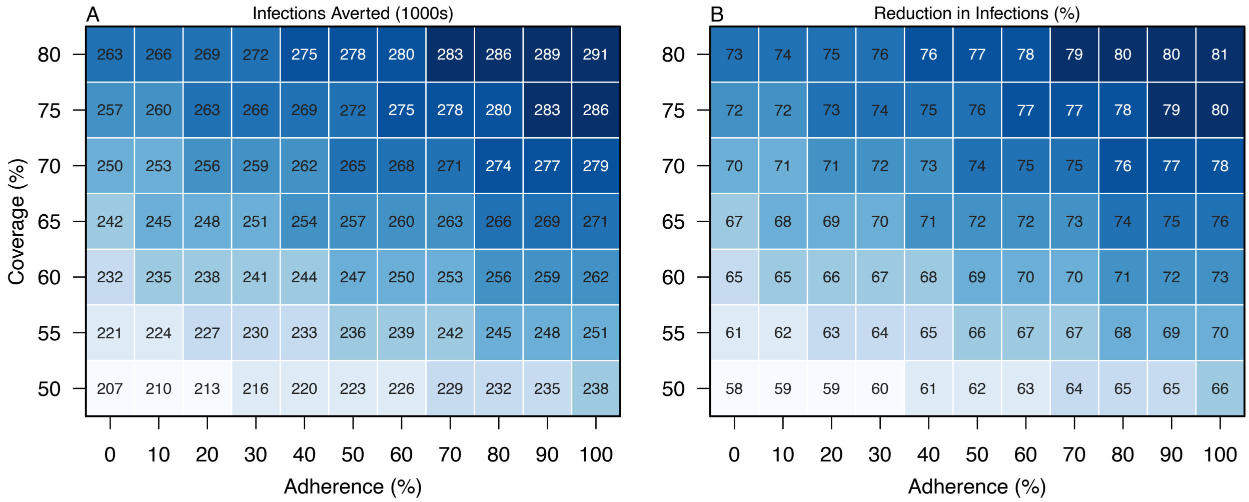


**Figure S16. Effect of coverage and adherence on the projected impact of MDA (CQ + PQ) for *P. vivax* in Sifontes**. The expected number of *P. vivax* infections averted (A) and the effect size (B) across 2021-2023 is presented for each combination of coverage and adherence in the target population. The intensity of the color denotes the median projected impact, where lighter colors denote lower impact and darker colors denote high impact.

**Uncertainty in LLIN Action**

We quantified the sensitivity of our LLIN impact projections to parameter values assumed in Table S6 by projecting the impact of LLINs under an alternative scenario in which the value of one parameter was 50% less than the assumed default value. For *P. falciparum* and *P. vivax*, the projected impact of LLINs was most sensitive to the half-life of LLIN usage and the mortality effect of the insecticide (Fig. S17). Under the assumption that half-life of LLIN usage was 50% less than the default value, we projected that 16,500 (95% PPI: 15,400 – 17,400) *P. falciparum* infections and 133,000 (132,000 – 134,000) *P. vivax* infections would be averted during 2021-2023 at a LLIN coverage of 10%, compared to 20,200 (18,700 – 21,200) *P. falciparum* infections and 168,000 (167,000 – 170,000) *P. vivax* infections under default assumptions. Similarly, under the assumption that mortality effect of the insecticide was instead 50% less than the default value, we projected that only 14,300 (13,300 – 15,000) *P. falciparum* infections and 113,000 (112,000 – 114,000) *P. vivax* infections would be averted during 2021-2023 at a LLIN coverage of 10%. That the LLIN projections are quite sensitive to the half-life of LLIN usage suggests that measures taken to increase the duration of LLIN usage among recipients would lead to a greater projected impact of the intervention.

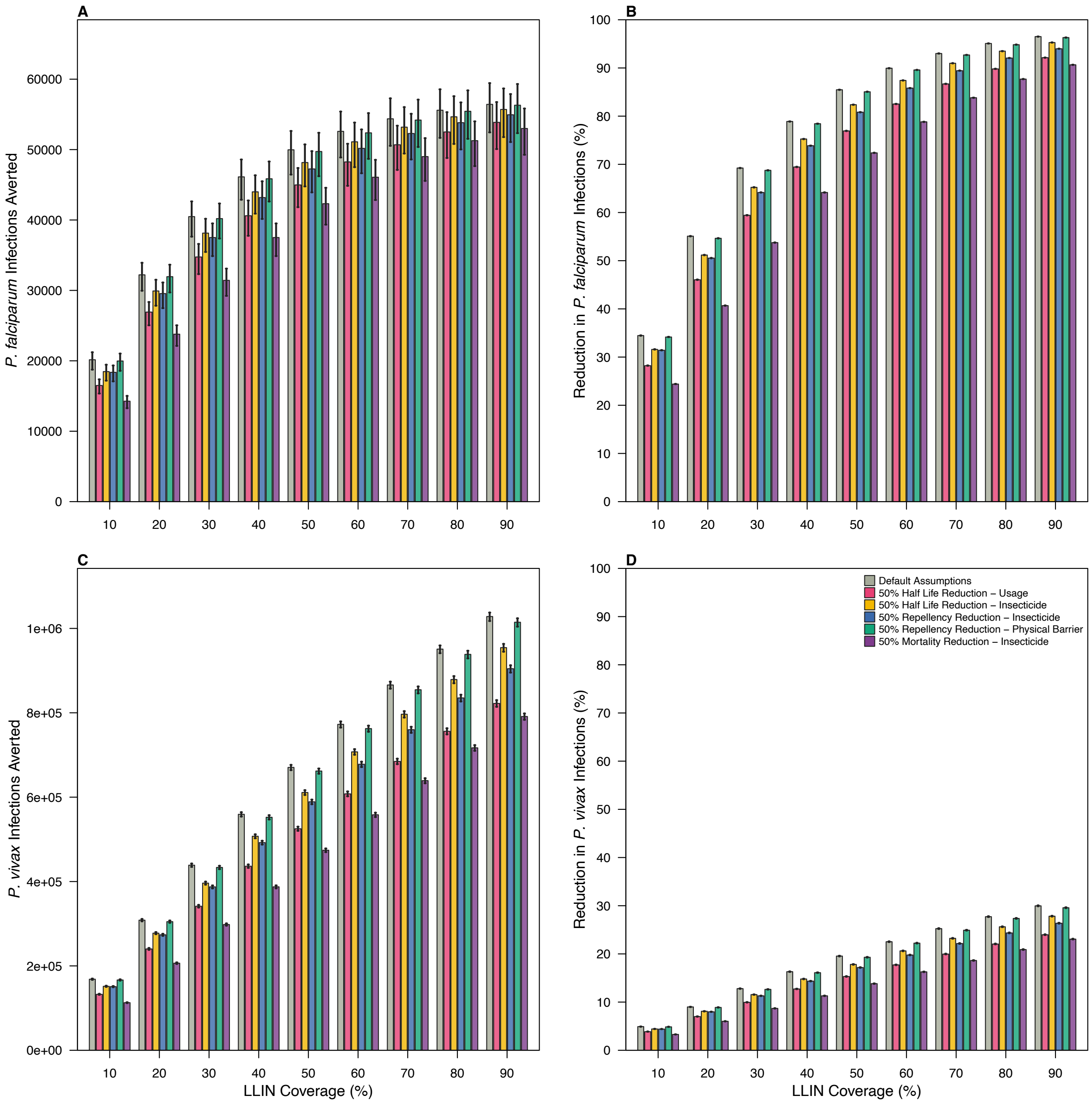


**Figure S17. Sensitivity of LLIN impact projections to model parameters.** The expected number of *P. falciparum* infections averted (A) and the percentage reduction in *P. falciparum* infections (B) during 2021-2023 and the expected number of *P. vivax* infections averted (C) and the percentage reduction in *P. vivax* infections (D) during 2021-2023 are presented as a function of coverage. Grey bars denote the impact projections made under the default assumptions presented in the main text, and all other scenarios measured the impact when each parameter of the LLIN intervention model was reduced by 50%. The parameters varied were the half-life of LLIN usage (pink), the half-life of the insecticide (yellow), the repellency effect due to the insecticide (blue), the repellency effect due to the physical barrier of the LLIN (green), and the mortality effect due to the insecticide (purple). The height of each bar is the median of the posterior predictions, and the vertical segments denote the 95% posterior prediction intervals.

**Sensitivity of Impact Projections to Inference Replicates**

Given that our Gelman-Rubin statistics did not provide support for convergence, we evaluated whether our intervention impact projections were sensitive to the lack of demonstrated convergence. We projected the impact of LLINs and MDA for each of the SMC chains and found that our results were robust to the choice of SMC chain

(Figs. S18-S20). These results indicated that the lack of formal convergence of the parameter estimates on the basis of the Gelman-Rubin statistic did not affect the quantitative nor qualitative conclusions that we reached about intervention impact.

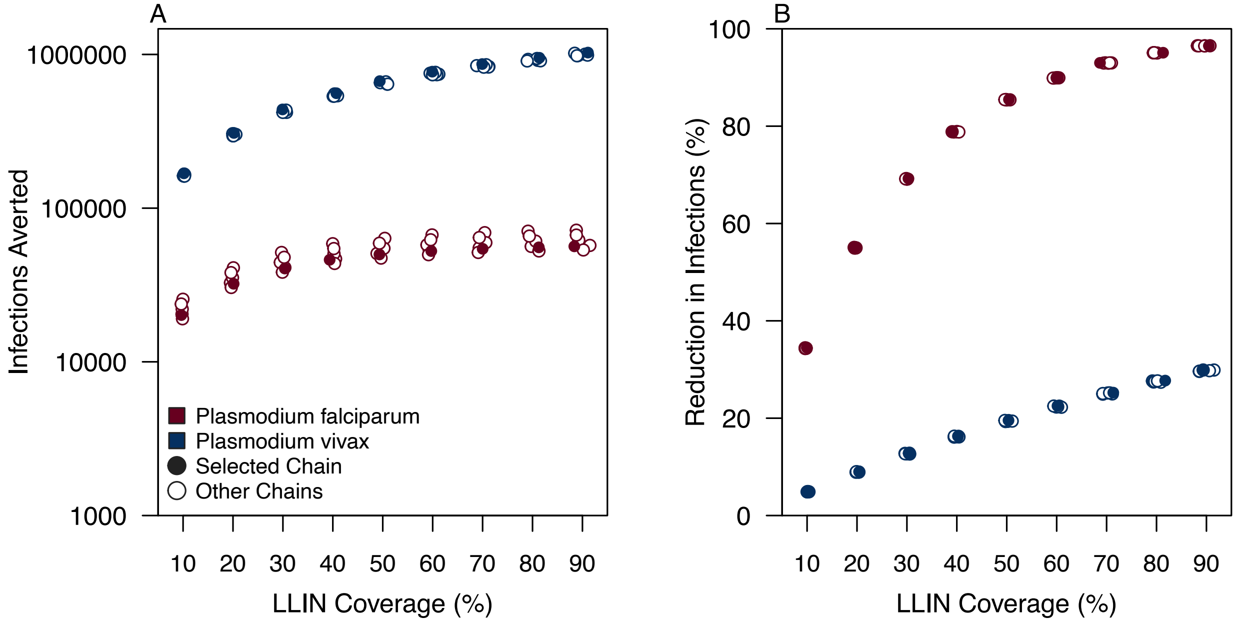


**Figure S18. Sensitivity of LLIN Impact Projections to Inference Replicates.** (A) The expected number of *P. falciparum* (red) and *P. vivax* (blue) infections averted during 2021-2023 is projected as a function of LLIN coverage for the inference replicate used in the primary analysis (closed cicle) and all other inference replicates (open circle). (B) The percent reduction in infections is projected as a function of LLIN coverage for all inference replicates. In both plots, the point denotes the median of the posterior prediction distribution, and the segments are the 95% posterior prediction interval.

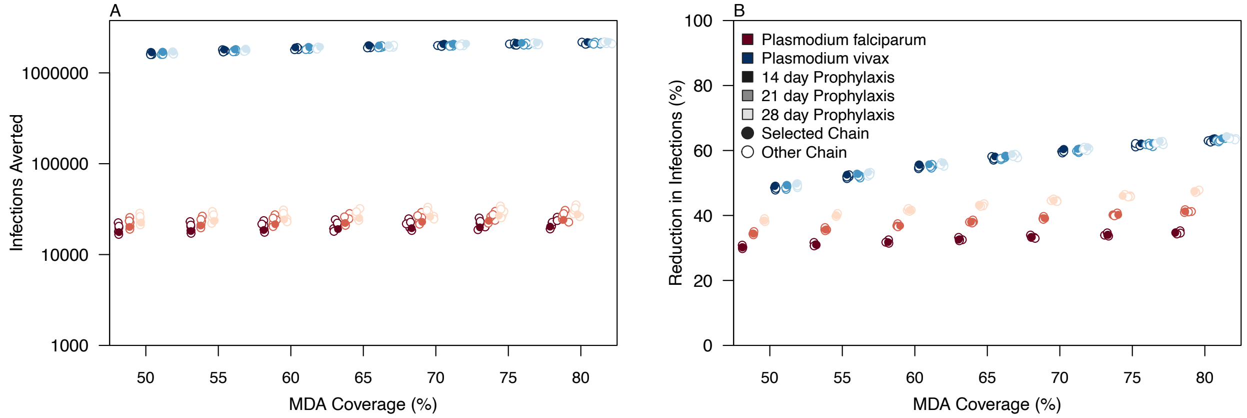


**Figure S19. Sensitivity of MDA (CQ vs. ACT) Impact Projections to Inference Replicates.** (A) The expected number of *P. falciparum* (red) and *P. vivax* (infections) averted during 2021-2023 is projected as a function of MDA coverage and varying durations of prophylaxis for the inference replicate used in the primary analysis (closed circle) and all other inference replicates (open circle). (B) The percent reduction in infections is projected as a function of MDA coverage for all inference replicates. In both plots, the point denotes the median of the posterior prediction distribution, and the segments are the 95% posterior prediction interval.

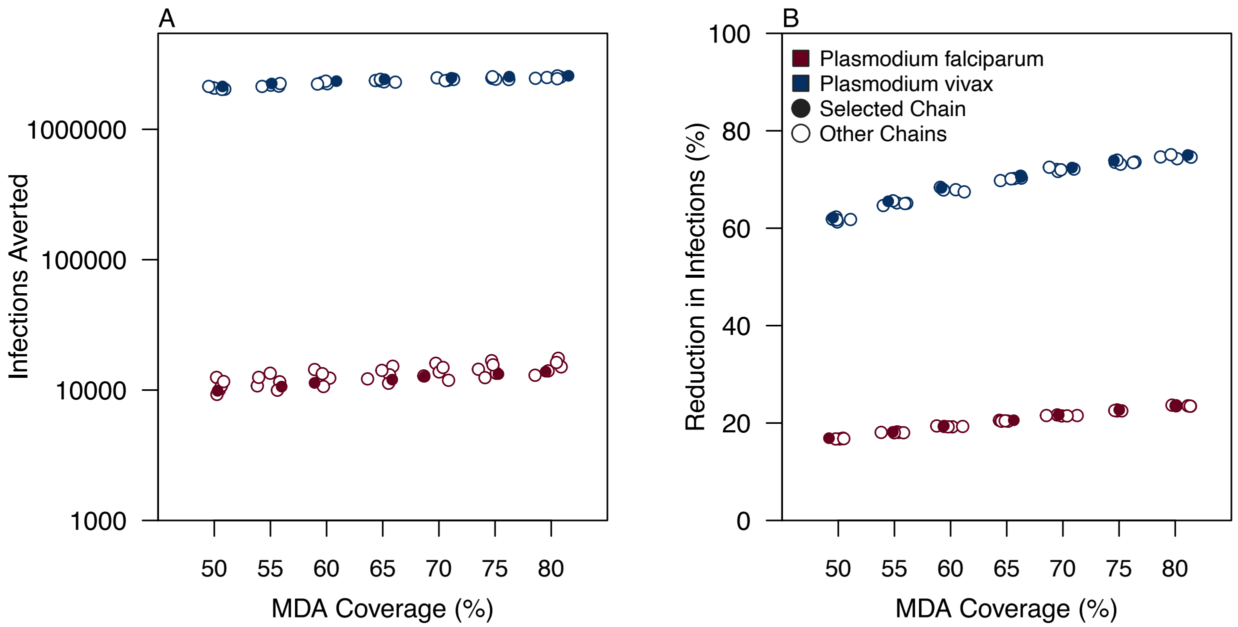


**Figure S20. Sensitivity of MDA (CQ + PQ) Impact Projections to Inference Replicates.** (A) The expected number of *P. falciparum* (red) and *P. vivax* (infections) averted during 2021-2023 is projected as a function of MDA coverage for the inference replicate used in the primary analysis (dark shade) and all other inference replicates (light shade). (B) The percent reduction in infections is projected as a function of MDA coverage for all inference replicates. In both plots, the point denotes the median of the posterior prediction distribution, and the segments are the 95% posterior prediction interval.
